# The local and systemic response to SARS-CoV-2 infection in children and adults

**DOI:** 10.1101/2021.03.09.21253012

**Authors:** Masahiro Yoshida, Kaylee B. Worlock, Ni Huang, Rik G.H. Lindeboom, Colin R. Butler, Natsuhiko Kumasaka, Cecilia Dominguez Conde, Lira Mamanova, Liam Bolt, Laura Richardson, Krzysztof Polanski, Elo Madissoon, Josephine L. Barnes, Jessica Allen-Hyttinen, Eliz Kilich, Brendan C. Jones, Angus de Wilton, Anna Wilbrey-Clark, Waradon Sungnak, J. Patrick Pett, Elena Prigmore, Henry Yung, Puja Mehta, Aarash Saleh, Anita Saigal, Vivian Chu, Jonathan M. Cohen, Clare Cane, Aikaterini Iordanidou, Soichi Shibuya, Ann-Kathrin Reuschl, A. Christine Argento, Richard G. Wunderink, Sean B. Smith, Taylor A. Poor, Catherine A. Gao, Jane E. Dematte, NU SCRIPT Study Investigators, Gary Reynolds, Muzlifah Haniffa, Georgina S. Bowyer, Matthew Coates, Menna R. Clatworthy, Fernando J. Calero-Nieto, Berthold Göttgens, Christopher O’Callaghan, Neil J. Sebire, Clare Jolly, Paolo de Coppi, Claire M. Smith, Alexander V. Misharin, Sam M. Janes, Sarah A. Teichmann, Marko Z. Nikolić, Kerstin B. Meyer

**Author notes:** co-first authors (Masahiro Yoshida, Kaylee B. Worlock, Ni Huang, Rik G.H. Lindeboom). co-senior authors (Marko Z. Nikolić, Kerstin B. Meyer).

## Abstract

While a substantial proportion of adults infected with SARS-CoV-2 progress to develop severe disease, children rarely manifest respiratory complications. Therefore, understanding differences in the local and systemic response to SARS-CoV-2 infection between children and adults may provide important clues about the pathogenesis of SARS-CoV-2 infection. To address this, we first generated a healthy reference multi-omics single cell data set from children (n=30) in whom we have profiled triple matched samples: nasal and tracheal brushings and PBMCs, where we track the developmental changes for 42 airway and 31 blood cell populations from infancy, through childhood to adolescence. This has revealed the presence of naive B and T lymphocytes in neonates and infants with a unique gene expression signature bearing hallmarks of innate immunity. We then contrast the healthy reference with equivalent data from severe paediatric and adult COVID-19 patients (total n=27), from the same three types of samples: upper and lower airways and blood. We found striking differences: children with COVID-19 as opposed to adults had a higher proportion of innate lymphoid and non-clonally expanded naive T cells in peripheral blood, and a limited interferon-response signature. In the airway epithelium, we found the highest viral load in goblet and ciliated cells and describe a novel inflammatory epithelial cell population. These cells represent a transitional regenerative state between secretory and ciliated cells; they were found in healthy children and were enriched in paediatric and adult COVID-19 patients. Epithelial cells display an antiviral and neutrophil-recruiting gene signature that is weaker in severe paediatric *versus* adult COVID-19. Our matched blood and airway samples allowed us to study the spatial dynamics of infection. Lastly, we provide a user-friendly interface for this data^1^ as a highly granular reference for the study of immune responses in airways and blood in children.

## Introduction

The severe acute respiratory syndrome coronavirus 2 (SARS-CoV-2) is responsible for the current coronavirus disease 2019 (COVID-19) pandemic. SARS-CoV-2 infection is relatively rare in children (1-2% of total cases reported <19 years)^2–4^, and generally presents with milder severity compared to adults^2, 3, 5, 6^. Hospitalisations, progression to symptomatic respiratory failure requiring oxygen due to SARS-CoV-2 pneumonia, ventilatory support in intensive care and death are all rare in children, with the most common clinical manifestation of the disease reported as fevers, cough, rhinorrhea, myalgia and fatigue^7–10^. Differences in disease progression between children and adults may hold clues for better treatment of severe SARS-CoV-2 infection.

SARS-CoV-2 employs a host cell surface protein, angiotensin-converting enzyme (ACE) 2, as a receptor for cellular entry^11^. A number of recent studies have suggested that the expression of *ACE2* is both tissue and age-dependent^12, 13^, and differences in *ACE2* expression between children and adults were proposed to contribute to less severe disease in children. At single cell level, viral entry genes were shown to be most highly expressed in the nasal epithelium in healthy adults^14^. In children, bulk RNA sequencing analysis revealed lower *ACE2* gene expression both in upper^15^ and lower airways^13, 16^ compared to adults, although more recent studies found no correlation with age or infection^17^.

Upon infection, 14% of symptomatic adults develop progressive respiratory failure displaying a strong inflammatory immune response^18^. Recent single cell analysis of this response demonstrated the involvement of various types of immune cells, including proinflammatory monocytes/macrophages^19–21^, clonally expanded cytotoxic T cells^22–24^ and neutrophils^22^. Although these specific cell types are beginning to be resolved in adults^19–21, 20, 22, 24, 25^, they have not been as comprehensively characterised in children. So far, only bulk RNA sequencing and cytokine studies have compared immune responses between children and adults, suggesting a more robust innate immune response, such as increased levels of interferon gamma (IFN-γ) and interleukin-17 (IL-17A) in plasma^26^, and a reduced antibody response and neutralising activity against SARS-CoV-2 in children^27^.

The immune landscape in early life has been shown to be distinct from that of adults^28–30^, in keeping with the immune system maturing from an immune tolerant state *in utero* to a more pro-inflammatory state with the increased exposure of new antigens and pathogens over the years^31^. Changes in blood cell counts and immune cell composition throughout childhood are well described^32^, and recently CyTOF (cytometry by time-of-flight) has facilitated a higher multiplexed panel-dependent description of cell types, with studies focussing on either early childhood^29^ or reporting comparison to disease^33^. Unbiased transcriptomic analysis of healthy paediatric blood immune cell types over the entire duration of childhood is lacking. Similarly, the upper airway mucosa only reaches maturity after puberty, in keeping with the high incidence of upper respiratory tract infections in children^34^, yet unbiased analysis of epithelial maturation is still missing.

To address these gaps in our knowledge and to identify paediatric-specific responses in COVID-19, we collected matched nasal, tracheal, bronchial and blood samples from patients across infancy, childhood, adolescence and adulthood and analysed them with single cell transcriptomics combined with protein profiling. We compare this with samples from paediatric and adult COVID-19 patients and report markedly different responses. In paediatric COVID-19, the biggest changes in blood are increased fractions of naive lymphocytes, regulatory T cells (Tregs) and innate lymphoid cells (ILCs) and a loss of natural killer (NK) cells and monocytes. In contrast, in the airways the proportion of activated macrophages, neutrophils and CD8^+^ T cells goes up whilst B lineage cells are underrepresented. In adult blood, we see expanded T clones which encompass distinct T cell subtypes, whilst children show less expanded T cell clones, particularly in COVID-19 patients. We also chart the complex relationship between immune cell types present in the blood and the nose of the same individuals.

Overall, the substantial differences in response to SARS-CoV-2 between children and adults reflect the changes of the immune landscape over developmental time, which in children are dominated by naive adaptive and innate cell responses, whilst those in adults are dominated by memory cell recall adaptive responses, illustrated by expanded lymphocyte clones. Lastly, we describe a novel epithelial cell type transitioning from secretory to ciliated cells that is present in COVID-19 patients as well as healthy children which, among other epithelial cell types, displays a striking antiviral and neutrophil-recruiting signature.

## Results

### Study cohort

We have assembled a cohort of 30 healthy children from the five World Health Organisation (WHO) age ranges - neonates (0-30 days; n=6), infants (1-24 months; n=6), young children (2-6 years; n=8), children (6-12 years; n=5), adolescents (12-18 years; n=5) - and profiled the cellular landscape in the upper airways (nasal and tracheal brushings) and matching peripheral blood mononuclear cells (PBMCs) from blood using single cell RNA sequencing (scRNA-seq) (**Figure 1a-b**). For one individual a bronchial sample was included that matched both nasal and blood samples. In addition, we sampled 4 paediatric COVID-19 patients spread across the age categories and 18 adult COVID-19 patients with a range of disease severities, with lower airways (bronchi) sampled in a subset (**Figure 1b**). Only patients who tested positive for SARS-CoV-2 by RT-qPCR were enrolled in the study, and symptom onset relative to RT-qPCR testing and sampling is summarised in **Extended Data Figure 1a**. For some patients blood was additionally sampled on the day of hospital discharge (labeled convalescent) and our cohort included patients who were recalled 3 months after recovering from COVID-19 requiring respiratory support at the time of infection (non-invasive or invasive ventilation), labeled as post-COVID (**Extended Data Figure 1a-b**). Patient characteristics and metadata are in **Extended Data Table 1a**. This includes ethnicity data, but we also used our data set to infer ethnicities and found that 48% of our cohort were of European, 16% of Asian, 10% of African and 26% of mixed descent (**Extended Data Figure 1c)**. Nasal, tracheal and bronchial brushings were freshly processed by cold digestion and analysed by Chromium 10X 5’ sequencing. Where possible, isolated PBMCs from matching patients were frozen, thawed, multiplexed and sequenced with 5’ 10X technology using a 192 antibody CITE-seq panel (**Figure 1b**; **Extended Data Table 2** for CITE-seq panel) generating matched transcriptional and protein data.

**Figure 1:**
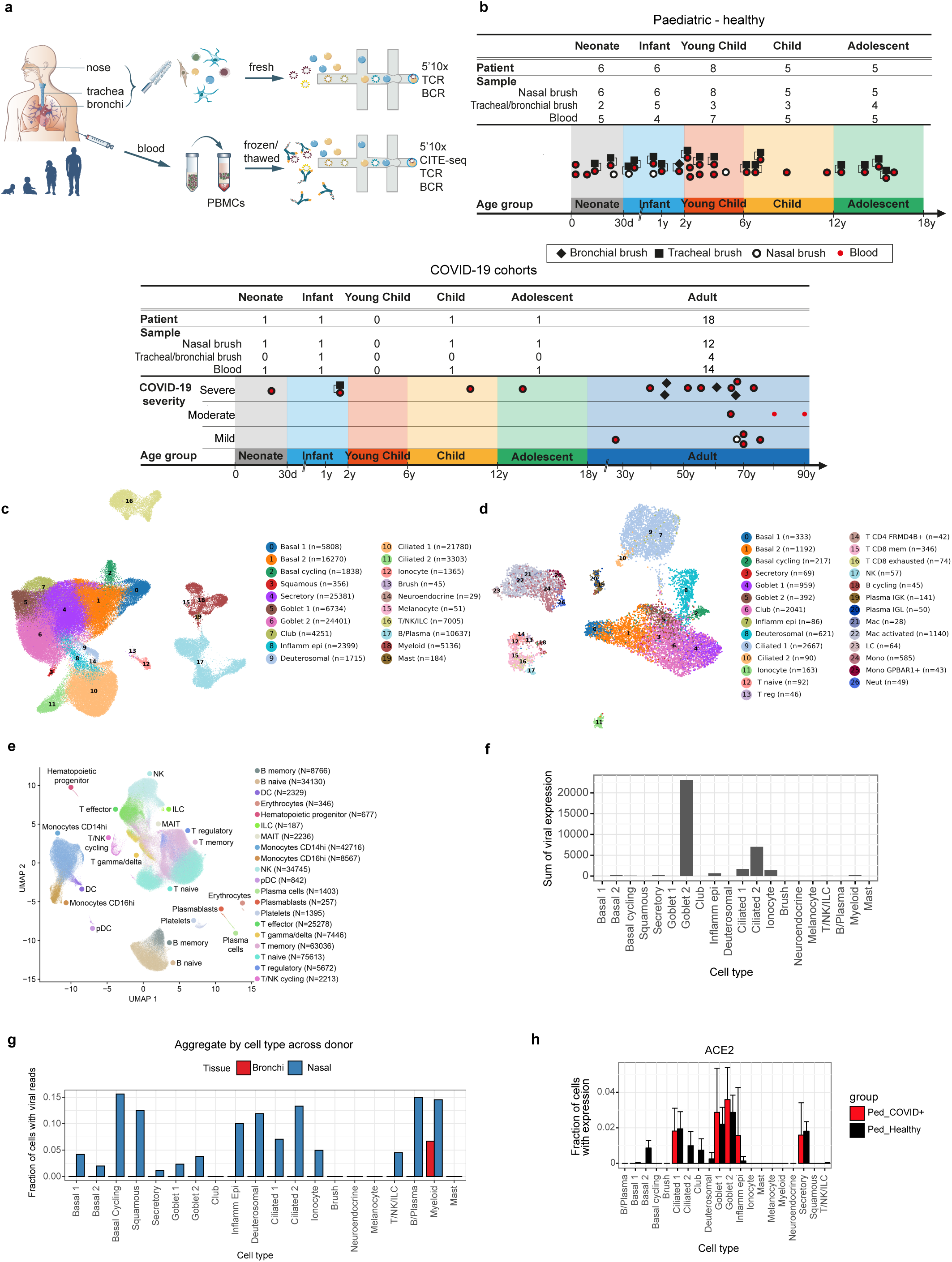
Experimental outline and overview of results. **(a)** Nasal, tracheal and bronchial brushings as well as peripheral blood were collected from across the human lifespan, from neonates to the elderly. The brushing samples were processed fresh for 10X 5’ single cell capture and sequencing, including TCR and BCR. PBMCs were isolated and then frozen for multiplexed processing later on. Upon thawing, PBMCs were stained with Total-seq-C reagents and processed for 10X 5’ single cell capture and sequencing, including TCR, BCR and CITE-seq. **(b).** Overview of samples taken in our healthy paediatric patient and COVID-19 cohorts. COVID-19 severity was classified as mild (symptomatic without oxygen requirement or respiratory support), moderate (requiring oxygen without respiratory support) or severe (requiring non-invasive or invasive ventilation). **(c)** UMAP visualisation of annotated upper airway (nose and trachea) cells, **(d)** and bronchial cells, with cell numbers per cell type in brackets. **(e)** UMAP visualization based on the RNA expression data of all PBMC cells annotated into broad blood cell types. **(f)** Total SARS-CoV-2 viral expression detected per cell type. **(g)** Fraction of cells with detected viral RNA molecules in different cell types and sampling locations. (**h**) *ACE2* expression in healthy and paediatric COVID-19 patients per cell type.

For the upper airways, we generated 138,688 high quality single cell transcriptomes. Of these 115,726 were epithelial and 22,962 immune cells (**Figure 1c, Extended Data Figure 2**). For the bronchi, a total of 11,672 cells were derived from 4 adult COVID-19 patients (**Figure 1d**). For PBMCs we obtained 317,854 cells of which 181,346 were isolated from COVID-19 patients (**Figure 1e**). The full dataset is available at https://www.covid19cellatlas.org/ for easy browsing and interactive data analysis.

### Detection of SARS-CoV-2 reads

When aligning transcriptomes, we included the SARS-CoV-2 reference genome, and detected viral reads (> 10 reads) in 4 individuals (3 nasal and 1 bronchial). In the upper airways, we detected the highest levels of total viral expression in the two most abundant cell types: goblet 2 and ciliated 2 cell populations (**Figure 1f**). When examining the percentage of cells with viral reads, fractions were similar across all nasal epithelial cell types (**Figure 1g**), which agrees with the expression of *ACE2* in our data set (**Extended data Figure 3a**). Viral reads were also detected in lymphocytes and myeloid cells (mostly macrophages), either reflecting active infection in macrophages^35^ or merely the uptake of virions or infected dead cells.

These findings are consistent with previous studies that reported highest levels of expression of *ACE2* and *TMPRSS2* in goblet and ciliated cells in healthy individuals^14^. Additional genes (*NRP1*^36, 37^, *BSG*^38^, *TFRC*^39^) have been implicated as secondary entry receptors (**Extended Data Figure 3a**) but in our data set of three individuals with high viral reads, only some correlate with actively infected cells (**Extended Data Figure 3b**). It has been reported that in adults, expression of *ACE2* is induced by interferon^40^ and in response to infection^25^. In contrast, we observe no significant increase of *ACE2* expression across cell types in children with COVID-19 (**Figure 1h**), consistent with recent bulk RNAseq comparisons^17^. However, we do see a difference in *ACE2* expression in adults, with higher expression in acute *versus* post-COVID patients (**Extended Data Figure 3c**). In line with previous reports^41, 42^, no viral reads were detected in peripheral blood within our COVID-19 cohort.

### Nasal, tracheal and bronchial epithelial cell analysis

We first focused our analysis on the epithelial cell compartment in the airways. Using unsupervised clustering and re-clustering (see methods) we detect 15 distinct epithelial populations (**Figure 2a**) based on known marker gene expression^43–45^ and indicate their distribution within the UMAP with respect to COVID-19 status and age (**Figure 2b**). In our cell type annotation, we used a conservative approach, using a high cut-off of 1000 UMIs per cell and normalising the data set by donor identity which ensures that cell type differences are consistent between individuals. We identified a small number of melanocytes, in keeping with previously reported primary nasal mucosal melanocytes^46, 47^. The major surface epithelial cell populations fall into two broad clusters, with the first covering the basal to secretory cell differentiation pathway, as visualised using Velocyto (**Figure 2c),** and these include basal 1, basal 2, cycling basal, secretory, goblet 1 and 2, and squamous cells with markers as specified in **Figure 2d**. As previously described^48^, basal cells express high levels of *TP63* and *KRT5* and we subdivided these by the expression of *KRT15* and *DLK2*, whilst basal 2 cells are high in *NOTCH1* and the early epithelial differentiation marker *DAPL1*. Secretory cells express a range of secretory proteins such as mucins and antimicrobial peptides and appear to be the differentiation intermediates that give rise to the more differentiated club cells that are marked by high expression of *SCGB1A1* and *SCGB3A1*. Goblet cells have high levels of *MUC5AC*, but can be further distinguished by the expression of *TFF1* in goblet 1 and *TFPI2* in goblet 2 cells (**Figure 2d**), with the latter expressing higher levels of innate immune genes and a distinct mucin profile as assessed by DEGs analysis. Furthermore, we detect squamous cells with the characteristic expression of *SPRR3* and *KRT78*. In addition to the unsupervised clustering, we visualised markers (*KRT14*, *KRT6A*, *KRT13*, *KRT24*) that have been associated with “Hillock” cells^44, 49, 50^ which seem to align along a distinct differentiation trajectory at the edge of the secretory cluster from basal towards squamous cells (**Extended Data Figure 4a**). Within the ciliated cluster, the differentiation trajectory points from ciliated 1 (*PIFO*, *OMG*) to ciliated 2 (*CFAP54*, *DZIP1L*) cells. Between the secretory and ciliated clusters, we observe deuterosomal cells as intermediates which are marked by *CDC20* and *FOXN4*^43, 44^ (**Figure 2c**). In addition, we also detect a novel, second cell population that forms a second bridge between the secretory and ciliated clusters. We refer to these as inflammatory epithelial transit cells (IETCs) as they are high in inflammatory markers such as *S100A8* and *S100A9*. They co-express ciliated cell markers such as *PIFO* and express secretory genes such as *MUC2*, but *FOXJ1* expression is low. Distinguishing markers are two long non-coding RNAs, FP671120.4 and FP236383.2. This population has not been described in previous analyses of adult airways^43, 44^. We detect these cells mostly in COVID-19 patients, but also in healthy children (**Figure 2b**), suggesting a function in development and tissue regeneration following injury (see below). We also detect rare cell types such as ionocytes, brush cells and neuroendocrine cells, each expressing their canonical markers^43, 44^.

**Figure 2:**
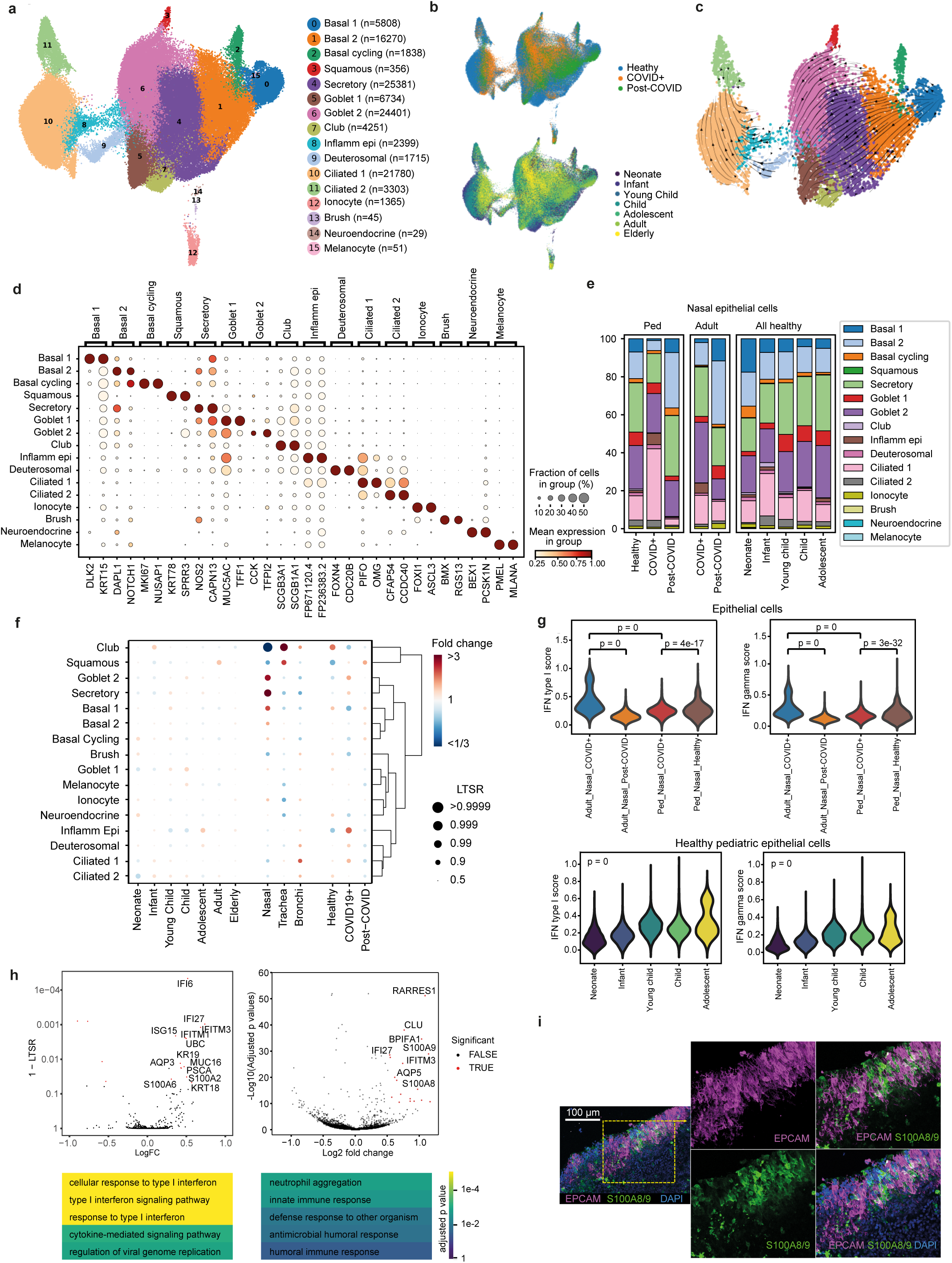
Epithelial cells. **(a)** UMAP visualisation of annotated airway epithelial cells. **(b)** Airway epithelial cells in the same UMAP as **(a)**, coloured by COVID-19 status and age group. **(c)** RNA velocity of major epithelial cell types. **(d)** Dot plot showing marker genes for annotated airway epithelial cell types, with fraction of expressing cells and average expression within each cell type indicated by dot size and colour, respectively. **(e)** Bar plot comparing epithelial cell type compositions across COVID-19 status and age groups. **(f)** Dot plot showing fold change and statistical significance of epithelial cell type proportions across age group, location of sampling and COVID-19 status, respectively, estimated by fitting Poisson generalised linear mixed models taking into account other technical and biological variables. (g) Comparing expression signature of cellular response to type 1 interferon (left) and interferon gamma (right) across COVID status in children and adult (top) and across age groups in healthy children (bottom). Wilcoxon ranked-sum test and Cuzick’s rank-sum test were used for pairwise comparisons and test of trend across age groups, respectively. **(h)** Differential gene expression contrasting COVID-19 and non-COVID-19 samples in inflammatory epithelial transit cells (IETCs) without (left) and with (right) age group controlled. Below each are top 5 enriched biological process GO terms in genes up-regulated in COVID-19 positive IETCs. **(i)** Confocal microscopy image illustrating expression of S100A9 (green) by epithelial cells (EpCam, magenta) in the nasal epithelium. Nuclei stained with DAPI (blue).

### Changes of epithelial cell type proportions in healthy and COVID-19 individuals

We next examined the epithelial cell type proportions in our cohort (**Figure 2e**,f). To test significance, we used a multivariate random effect model (see methods) that allowed us to take into account clinical metadata, such as age, sex, ethnicity, tissue, COVID-19 status and six technical factors (**Figure 2f**), of which donor and tissue accounted for the largest fraction of variance (**Extended Data Figure 4b**). Airway epithelial cell type composition does not change extensively with age (**Figure 2f, Extended Data Figure 4c**) but notably with location, with an increase in goblet 2 and secretory cells in the nose compared to bronchi and trachea. Conversely, there is a decrease in club cells in the nose. Statistical significance is indicated by local true sign rate (LTSR), which was greater than 0.999 for the latter observation.

Contrasting COVID-19 *versus* healthy patients, the most highly enriched cell types are inflammatory epithelial transit cells (IETCs), goblet 2 and ciliated 1 cells. In contrast, basal 1 cells are notably reduced. Ciliated and goblet cells contain the highest number of viral reads (**Figure 1f**), most likely leading to cell death^50, 51^. We hypothesize that increased ciliated 1 and goblet 2 cell numbers reflect an ongoing compensatory replacement of these dying cells by their precursors to maintain homeostasis as seen in lower airways upon infection^52, 53^. Differentiation trajectory analysis suggests that the IETCs serve as an alternative differentiation path for repopulation of ciliated cells (**Figure 2c**), leading to a depletion of basal 1 cells, the main stem cell population within the airway epithelium. This is further supported by the finding that in post-COVID patients the basal 1 population recovers (**Figure 2f**).

### Weak interferon response in children with severe COVID-19

In adults viral infection is strongly associated with an interferon response^54^ and for SARS-CoV-2 a higher response to interferon has been reported for mild and moderate disease than for severe disease^50, 55, 56^. We therefore carried out differential gene expression analysis (using a multivariate random effect model; see methods) between healthy controls and COVID-19 patients, followed by a GO term enrichment analysis. For the strongly enriched inflammatory epithelial transit cells (IETCs) the DE genes in healthy *versus* diseased patients were enriched for interferon responsive genes (**Figure 2h**) and were associated with an interferon type I, antiviral and cytokine response. When testing interferon-alpha and gamma response signatures across epithelial cell types, expression was highest in secretory and goblet cells (**Extended Data Figure 5a**). Healthy children showed an interferon response that increased with age (**Figure 2g**). A comparison across conditions (**Figure 2g**) showed that the interferon response in adult COVID-19 patients was higher than in children with COVID-19. As recent bulk RNAseq data suggest that interferon responses correlate with viral load, but not age^17^, we interpret this to indicate that like in adults, severe COVID-19 disease in children is associated with reduced interferon responses.

### A neutrophil recruiting gene signature associated with COVID-19

In addition to the above analysis, we also compared gene expression in inflammatory epithelial transit cells (IETCs) without correcting for age (**Figure 2h**) and saw upregulation of innate immune response genes with high expression of serum amyloid protein (*SAA1*), an acute phase protein highly expressed in inflammation and tissue injury^57^, and *S100A8* and *S100A9* which together encode calprotectin. Calprotectin is also highly expressed in myeloid cells and has been identified as a key mediator of mild *versus* severe COVID-19^58^. Using this gene set in GO term analysis highlights neutrophil enrichment as the top enrichment term (**Figure 2h**), a striking finding given that neutrophil infiltration is linked to COVID-19 disease severity^59^. As calprotectin expression has primarily been associated with myeloid cells, we wished to validate expression at the protein level in epithelial cells. **Figure 2i** depicts multiple dual positive cells, staining for both calprotectin and the epithelial marker EPCAM in a sample taken from the posterior nasal space of an adult COVID-19 patient. At the RNA level, calprotectin is expressed across different secretory cell types (**Extended Data Figure 6**).

### Comparison to transitional populations in different locations

Previous studies of adult nasopharyngeal swabs had found a population of interferon responsive cells (IRC), as an intermediate between secretory and ciliated cells^25^. We therefore compared the “transitioning” populations in the Chua *et al* data set with those in our upper airway (nose and trachea) and bronchial cells (**Extended Data Figure 6)**. All locations contained equivalent deuterosomal cells. The IRC makers *ISG15*, *IFIT1* and *CXCL10* were widely expressed in all three data sets, but only associated with transitioning cells in the Chua *et al* data set. The inflammatory epithelial transit cell (IETC) markers *SAA1*, *S100A8* and *S100A9* were found in transitioning cells in both upper airway data sets, but less obviously in transitioning populations in the bronchial data set (**Extended Data Figure 6**). Thus, transitioning cells are likely to be plastic in their phenotype^48^ with exact gene expression signatures depending on the site of sampling, the interval between disease onset and sampling and other clinical covariates.

### Immune cell states in healthy paediatric blood defined by multi-omic data

Next we examined the cellular landscape of the healthy developing immune system in both blood and the upper airways in children. We generated CITE-seq data for 51 blood samples using a 192 antibody panel recognising 188 proteins. By integrating gene and protein expression data using an integrated Weighted Nearest Neighbor analysis^60^ and Leiden clustering^61^, we were able to annotate our 317,854 single cells into 31 cell types (**Figure 3a**, quality control metrics in **Extended Data Figure 7**). While the gene expression data made a greater contribution to distinguishing cell types, the protein expression data strongly contributed to defining T cell subtypes and monocytes, as visualised by pie charts in **Figure 3a**. Notable genes for which protein expression was more informative than RNA expression levels included CD4 in T cell subsets, NCR1 and CD56 (NCAM1) in NK cells, and CCR6 and IGHD in naive and memory B cells (**Figure 3c**, **Extended Data Figure 8a**). CITE-seq also allowed us to distinguish between cell type-specific protein isoforms, including CD45RA and CD45RO, which are key markers of T cell maturity. While RNA marker genes were consistent with previous reports^24^, the relative contribution of protein data varied significantly between cell types (**Figure 3a, Extended Data Figure 8a**). For example, for T cell subsets transcriptional data carried only slightly higher weight than the protein data, whilst plasmablasts, cycling T and NK, and haematopoietic stem and progenitor cells (HSPCs) were almost entirely assigned on the basis of RNA expression. To validate our manual cell type annotation and assess its accuracy, we compared it to an automated annotation by Azimuth^62^. This revealed that our annotation was highly similar, but not identical to automated annotation (**Extended Data Figure 8b**).

**Figure 3:**
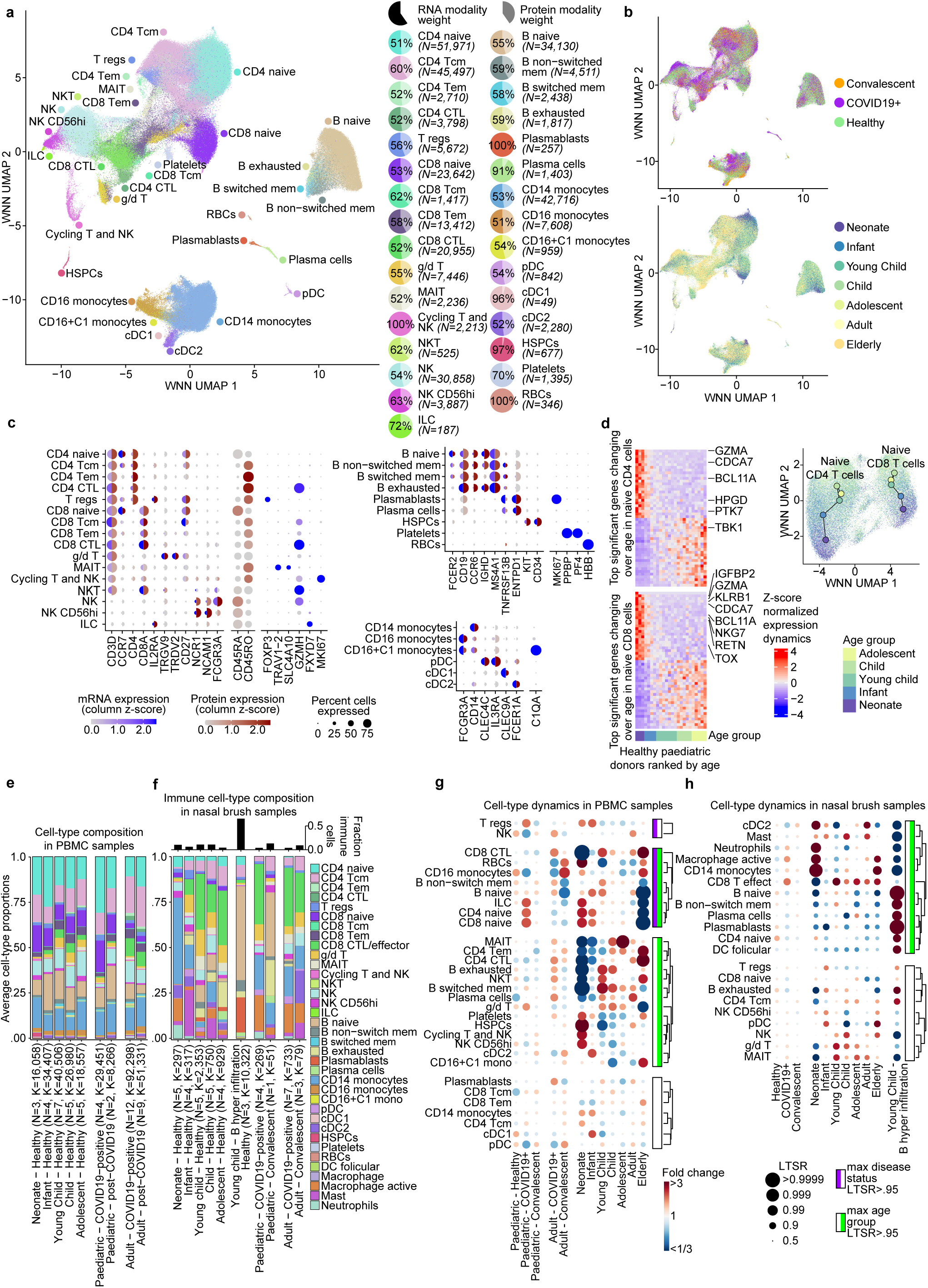
Immune analyses. **(a)** UMAP visualization of PMBCs based on integrated RNA and surface protein expression. UMAP was derived from a WNN graph that incorporates both protein and RNA expression data. The modality weights of the RNA and protein data are shown as pie charts in the legend. Cell types were annotated based on semi-supervised Leiden clustering and marker expression. **(b)** UMAPs as in **(a)** in which the COVID-19 status (top panel) and the age group (bottom panel) is visualized for each cell. **(c)** Dotplot of matched RNA expression (blue half) and surface protein expression (red half) of cell type markers. The size of the dot is scaled to the percentage of cells that have at least one count of each gene or protein. The color is scaled to the z-score normalized expression of each gene or protein compared to all other cell types (see **Extended Date Figure 8a**). **(d)** Heatmap showing the relative expression dynamics of the top 50 most changing genes that significantly change over age (FDR<0.001) within naive CD4 T cells (top heatmap) and within naive CD8 T cells. Genes that are known to be related to T cell identity are highlighted. The inset UMAP visualizes how naïve T cells from different age groups separate after reselecting hypervariable genes within this subset. This panel only includes data from healthy paediatric donors. **(e-f)** Barplots showing the average immune cell type proportions in PBMC samples (in **e**) and in nasal brushing samples (in **f**). Cell types are color coded as shown in **f**. Samples are grouped on age and COVID-19 status. *N* denotes the amount of samples in each group, while *K* denotes the amount of cells per group. Three healthy young children with very high B cell counts in their nasal samples were identified as outliers (see **Extended Data Figure 9c)** and plotted separately. In **f**, the fraction of immune cells in all cells identified in the nasal brushing samples are quantified on top. **(g-h)** Dot plot showing fold change and statistical significance of epithelial cell type proportions across age group, location of sampling and COVID-19 status, respectively, estimated by fitting Poisson generalised linear mixed models taking into account other technical and biological variables. The cell types that change significantly in at least one age and COVID-19 status group, are split and highlighted in green and purple, respectively. **(a-c,e-h)** Tcm, T central memory; Tem, T effector memory; CTL, cytotoxic T lymphocyte; T regs, regulatory T; g/d T, gamma/delta T cells; MAIT, mucosal-associated invariant T; NK, natural killer; NKT, natural killer T; ILC, innate lymphoid cells; pDC, plasmacytoid dendritic cell; cDC, conventional dendritic cell; HSPCs, hematopoietic stem and progenitor cells; RBCs, red blood cells. In these figures convalescent nasal brush samples refers to post-COVID airway samples only, with post-COVID and convalescent blood samples included in convalescent PBMCs.

Our data set allowed us to track changes in gene expression within cell types with age. In particular we noticed that both naive CD4^+^ and CD8^+^ T cells displayed a developmental pattern of gene expression (**Figure 3b,d**). A number of genes expressed in the neonatal age group were associated with cell proliferation or cancer (e.g. *CDCA7*, *BCL11A*), possibly reflecting a higher proliferative capacity in these early cells. The presence of *GZMA* (granzyme A) in both CD4^+^ and CD8^+^ naive T cells and *NKG7* (Natural killer cell granule protein 7) in only the CD8^+^ cells may suggest that these early naive T cells have specialised immune functions, with potentially higher cytotoxic activity. Other genes regulated over developmental time included those with potential immune regulatory functions such as *HPGD* and *TOX*, with possible roles in Treg and T cell development, respectively^63, 64^.

We also observed age-related gene expression gradients in naive B cells and to a lesser extent in monocytes (**Figure 3b**) and carried out a similar trajectory analysis (**Extended Data Figure 8c**). There were clear changes observed for B cells, but less obvious changes for monocytes. Genes with changing expression over developmental time in naive B cells have functions connected to differentiation, regulation, glycosylation and cell adhesion (including *ILR4*, *KLF9*, *ADAM28*, *HMGA1*, *LARGE1* and *ITM2C*). In addition, markers that point to a more innate-like type of immune response, such as *CD9* and *CD1c*, which are associated with the marginal zone (MZ) B cell subtype^65^, are expressed at a higher level in neonates. Such changes in the characteristics of circulating B cells at a very young age might also be linked to the formation of structures like the marginal zone, which is not fully developed until 2 years of age^66^. Together our data suggests that naive T and B cells that have already entered circulation continue to develop and mature during the first year after birth.

### Immune cell proportions in blood and nose throughout childhood and in adults

We further investigated changes in immune cell type proportion over childhood and in disease in blood and nose (**Figure 3e-h**), using the same statistical model as described above. In this analysis all samples were considered simultaneously while accounting for technical variation, and effects for age and COVID-19 status were plotted for the paediatric and the adult cohort (**Figure 3g,h, Extended Data Figure 9a**). Changes that are associated specifically with disease, age or both are highlighted. We first focused on age related changes in PBMCs: in healthy neonates, we observed high proportions of HSPCs, however this was driven by a single individual (**Figure 3g**). Fractions of ‘Cycling T and NK’, ILCs and NK CD56hi were also high in neonates but reduced in infants. Levels of RBCs and platelets are shown but may be confounded by technical variation. Relative levels of naive CD8^+^ and CD4^+^ T cells and Tregs were high in neonates and infants and then decreased with age, with very low levels in the elderly (adults >65 years), consistent with ongoing immune stimulation over the life span. Proportions of conventional dendritic cells (cDCs) type 1 and 2, Tregs, and naive B cells increased from neonates to infants, but then decreased with age. In contrast cell types associated with an adaptive immune response such as CD8^+^ cytotoxic T lymphocytes (CTL), plasma cells and gamma delta (*γ*/*δ*) T cells all showed a striking increase in young children (2 to 6 years of age), presumably reflecting greater antigen exposure of children in this age group, for example in pre-school nurseries. Non-switched, switched and exhausted B cells were highest in young children and children. At the other end of childhood, mucosal-associated invariant T (MAIT) cells were increased in children and adolescents. The trend of decreased naive and increased adaptive immune cells continued in adults (**Figure 3g**).

### Changes in cell type proportions in blood in COVID-19

We next compared the relative cell type proportions in relation to COVID-19 (**Figure 3b**). Strikingly and in contrast to adult COVID-19, in paediatric patients with severe COVID-19 the proportion of naive B or T cells, Tregs and ILCs is increased (**Figure 3e,g**), on a background of little change in the average lymphocyte counts (**Extended Data Table1**), consistent with a previous report that COVID-associated lymphopaenia is less pronounced in children^67–69^. High numbers of naive cells may be due to increased release of immature B and T lymphocytes from the bone marrow and thymus respectively, or due to migration of more mature cells to the site of infection (also see **Figure 3e-h**). As expected, in convalescent paediatric COVID-19 the increase in naive cells was reversed. In addition, we see a decrease in monocytes and NK cells, possibly reflecting migration of these cells to the site of infection. Relative to children, our adult COVID-19 cohort displayed increased levels of adaptive immune cells, such as CD8^+^ CTL, CD4^+^ CTL, plasma cells and plasmablasts as well as exhausted B cells. The changes in cell type proportions reported here for adults include a comparison to paediatric samples, and would therefore not necessarily reflect the changes seen when comparing only healthy to diseased adults, where lymphopaenia and an increase in monocytes have been reported^70^. We also examined the interferon response in blood, which was highest in monocytes, and was found to be lower in the paediatric than the adult COVID-19 cohort (**Extended Data Figure 5b,c**). Taken together, we see large differences in SARS-CoV-2 immune responses between children and adults. Some changes, such as in Tregs, are disease specific, whereas others (naive T lymphocytes, monocytes) are associated with both disease and age or age only (e.g. increases in cytotoxic T cells). The response to SARS-CoV-2 in adults *versus* children thus strongly reflects the underlying changes in the immune landscape that we observe over the life span.

### The immune landscape in the upper airways with age and COVID-19

When investigating the immune cell proportions in the nose (**Figure 3f**) we again observed significant changes over the life span. Because the total number of immune cells retrieved in airway samples was far lower than for blood and no protein expression data was available, we used a broader T effector cell type annotation. Nevertheless, we annotated 21 different immune cell types and detected tissue resident populations such as follicular dendritic cells, macrophages, activated macrophages, neutrophils and mast cells. Cell surface markers used for classification are consistent with previous studies (**Extended Data Figure 9b**). Nasal brushings harvested both non-immune and immune cells and proportions varied between patient groups (**Figure 3f, Extended Data Figure 9c**). In our data set, we noticed three young children with a very high proportion of immune cells, possibly reflecting non-symptomatic respiratory infections that are likely to be common in this age group, and treated these patients as a separate subgroup.

When examining cell type proportions in the nose with age, the pattern of cell type distribution differed greatly between the airways and the blood. In the upper airways, healthy neonates had high proportions of activated macrophages, CD14^+^ monocytes, cDC2 and neutrophils that rapidly decreased with age. Young children had higher levels of CD8^+^ T effector, *γ*/*δ* and MAIT cells, that remained relatively high throughout childhood. Within the airway samples, a comparison of adolescents to adults and elderly revealed relatively few striking changes. Amongst the children with high immune infiltrates there was a very strong overrepresentation of B cell lineages (naive B, B exhausted, plasmablasts and plasma cells), follicular dendritic cells as well as naive T cells, likely reflecting the different phases of an acute adaptive immune response.

When comparing COVID-19 to healthy cells, we first examined differences separately, within the paediatric cohort and within the adult cohort. Mostly, changes were similar and we therefore aggregated the two data sets. The combined analysis showed that in the airways COVID-19 samples were enriched for activated macrophages, neutrophils and CD8^+^ T effector cells (**Figure 3h**). These changes are consistent with those described in other studies^20, 25^, but strikingly different to the changes we observe in the same individuals in blood (see below).

### Correlation of immune cell types across nose and blood

In our data set, we have the unique opportunity to compare cell type proportions between the upper airways and blood from the same individuals and assess how the relationship between these compartments changes over childhood and with disease. We first integrated the immune cell data set from the different sites (**Figure 4a**) and then examined how the cell type proportions change with disease or age (within the healthy paediatric data set) across the nose and blood (**Figure 4b**). As not all cell types were represented in both data sets labels on the x and y axis are not symmetrical. In healthy children, samples with highest T effector cells, MAIT and *γ*/*δ* T cells in PBMCs also contained high levels of analogous cell types in the airways. Monocytes and NK cells also correlated across the two sites, but tended to be lower when lymphocyte proportions were increased, almost falling into a checker-board pattern by unsupervised clustering. This contrasted sharply with the results in adult acute disease, where different lymphocyte sub-populations varied strongly in blood and no longer correlated uniformly with the T cell subsets in the upper airways. For example, MAIT cells and CD4^+^ central memory T (Tcm) cells, defined in both compartments, now show anticorrelation between the two sites. A significant decrease in circulating MAIT cells, paired with an increase in activation markers and infiltration in airways and lungs, has been observed in adults with an active infection^71–74^, suggestive of migration of MAIT cells to the airway epithelium as part of the response against SARS-CoV-2 infection.

**Figure 4:**
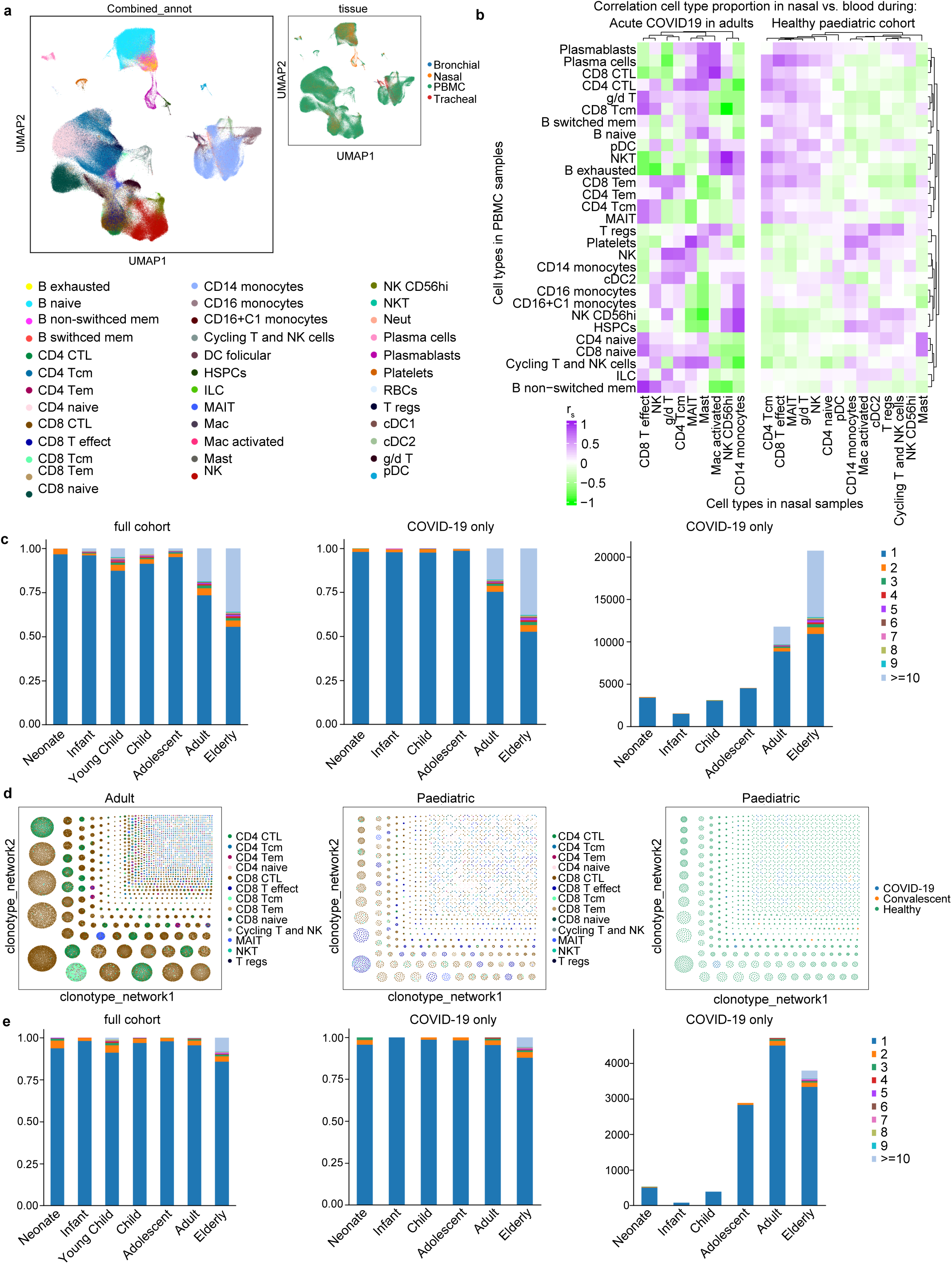
Immune cell types across compartments. **(a)** UMAP visualisation of the combined blood and airway data set, annotated by cell type and by site of sampling. **(b)** Correlation heatmap of the immune cell type proportions in the blood and nose within individuals. Spearman’s rank correlation coefficient between the relative cell type proportions in the blood and nose were calculated within all adult COVID-19 patients (left heatmap) and within all healthy paediatric samples (right heatmap). Both heatmaps were ordered using hierarchical clustering on the combined heatmaps. We considered only cell types that were present in at least half of the samples for correlation analysis. **(c)** Bar chart depicting the fraction of T cells clones (with single pair TCR*α*/*β* chains) detected within the full combined scRNA-seq dataset (left) and for the COVID-19 positive cohort only, shown as a fraction (middle) and total number (right). Clone size is indicated by colour and group by age. **(d)** Adult (left) and paediatric (middle) T cell clonotype networks coloured by cell type as well as by paediatric COVID-19 status. **(e)** Fraction of B cells clones (with single pair chains) detected within the combined scRNA-seq dataset (left) and the COVID-19 positive cohort only, shown as a fraction (middle) and total number (right). Note: No COVID-19 positive young children were collected so this age category is missing in **c** and **e**.

Overall, our findings highlight the need to study the sites of infection such as the upper airways as well as blood in order to fully understand the immune response to infections.

### Lymphocyte Clonality

From our 5’ 10X single cell libraries we amplified T cell receptor (TCR) and B cell receptor (BCR) sequences to analyse clonality. We analysed the VDJ data using scirpy (see methods) and found productive single pair TCR*α*/*β* chain rearrangements for all T cell subtypes, but also noted cells without productive TCRs (**Extended Data Figure 10a-b)**. The largest number of expanded T cell clones derived from peripheral blood, but we found the greatest proportion of large clones (>10) in the nose, with trachea also showing evidence of T cell clonal expansion (**Extended Data Figure 10c)**. When examining clone size across age, there was a consistent increase in expanded clones in older individuals (**Figure 4c**): in neonates and infants few expanded clones were observed, which increased in healthy young children and children, consistent with the evidence of stimulation of the adaptive immune system at that stage of childhood which we describe above (**Figure 3e,f**). Over adult life the number of large clones continued to increase, with the greatest expansion observed in the elderly (**Figure 4c**). We saw no identical clones between individuals, but we do see shared clones in different samples from the same individual (**Extended Data Figure 10d**), with most large clones coming from blood. In line with previous reports, we found that in adults the most highly expanded clones are CD8^+^ CTLs and CD8^+^ effector memory T (Tem) cells^75^ as well as CD4^+^ CTLs (**Figure 4d**). Consistent with expectations we see shared TCRs between CD8^+^ CTLs and CD8^+^ Tem and CD8^+^ Tcm; and separately between CD4^+^ CTL and CD4^+^ Tcm. In comparison to adults, we note that in the paediatric data individual clones were much smaller, and that in our cohort of patients the majority of expanded clones were from healthy children. Expanded clones were mostly CD8^+^ CTL and CD8^+^ T effector cells with fewer CD8^+^ Tem or CD8^+^ Tcm cell clones (**Figure 4d**). Low level of clonal expansion seen in COVID-19 paediatric patients could be due to overrepresentation of naive T cells, which show minimal clonal expansion (**Figure 4d-f**), and underrepresentation of CD8^+^ CTLs in paediatric PBMCs (**Figure 3e**). However, there is no evidence of defective activation as the proportions of nasal CD8^+^ CTLs in COVID-19 children *versus* adults are similar (**Figure 3e**).

We also studied expansion of B cell clones (see methods) (**Extended Data Figure 10 e-h**) and observed a very similar pattern. The number of expanded B cell clones increased with age and for the severe COVID-19 paediatric patients we saw fewer expanded clones than in the total cohort that included healthy children (**Figure 4e**).

In conclusion, we observed much greater clonal expansion of T and B cells in adult compared to paediatric patients, with a greater frequency of expanded memory and effector cells in adults. These observations further characterise the differences in age-specific immune response to COVID-19, most likely determined by the differences in the underlying immune landscape.

## Discussion

The analysis of our multi-omic single cell dataset of matched airway epithelial and blood immune samples in healthy and COVID-19 affected children and adults provides several significant advances in our understanding of normal immune and airway development, as well as COVID-19 pathogenesis. We present a detailed study of the immune and epithelial landscapes over healthy childhood development at single cell resolution. We used this to show clear differences between the immune and airway epithelial response to SARS-CoV-2 between children and adults, while also comparing the immune response in blood and airways in the same individuals. Additionally, we identify a novel epithelial cell state which is seen in COVID-19 patients and healthy children.

Our key findings in the healthy blood of children is the observation of unique gene expression signatures of naive B and T cells in the first year of life, that show hallmarks of innate immune responses^76^. These gene expression signatures are also present in fetal lung (unpublished observation) indicating that a number of fetal characteristics of the immune system persist in infancy.

A key motivation for our analysis was the current COVID-19 pandemic, which frequently starts with infection of the upper airways, where we found the highest total viral load in the surface epithelial goblet and ciliated cells. Viral infections are cleared by cell death and removal of infected cells^77, 78^, which led to a highly dynamic re-structuring of the airway epithelium with a marked increase in developmental intermediates, most notably the novel inflammatory epithelial transit cell (IETC) population, which was not found in previous analysis of healthy adult airways^25, 43, 44^, and a decrease in basal progenitors that is re-balanced post-infection. Upon infection, all adult epithelial cell types showed a strong interferon response, which was markedly reduced in our paediatric patients. Further analysis of mild paediatric COVID-19 cases will have to determine whether this characterises all responses in children or is limited to severe disease. The lack of interferon signals in the airways may explain why ACE2 expression is not induced in paediatric COVID-19. In our data set we also see a strong neutrophil recruiting signature, driven by expression of calprotectin in different epithelial cell types, thus highlighting the key role of epithelial cells in initiating an innate immune response.

The major distinguishing feature of the immune response to SARS-CoV-2 in children in blood are striking increases in naive B and T cells as well as ILCs, with a loss of CD16 monocytes. The lymphocyte response is strikingly different to COVID-19 immune responses in adults, described here and by others^23, 24, 56^, characterised by general lymphopaenia, but with increased CD8^+^ effector cells, as well as proliferating CD4^+^ and CD8^+^ cells in more severe disease. Whilst activated B lineage cells are reduced in paediatric COVID-19 patients, adult patients show increased plasma cells and plasmablasts, reported to be mainly the IgG subtype, in severe disease^24^. These differences appear to have their origin primarily in the different structure of the immune repertoire in children and adults that is further discussed below. Another difference we found between children and adults is the upregulation of Tregs and downregulation of NK cells in children compared to adults.

A unique aspect of our study is the ability to relate changes in the blood to those in the airways. In healthy children the proportions of immune cell types largely correlate between nose and blood. However, during active infection this relationship breaks down, highlighting the need to study both the active site of primary infection as well as blood to understand the dynamics of infection. For example, high levels of CTL CD8^+^ and MAIT cells in the nose are correlated with a reduction in the blood, likely reflecting the migration of these cells to the site of infection.

We also examined clonal expansion of B and T cells in our data set. We find that adults have far greater numbers of large, expanded T and B cell clones than children who show greater immune diversity. The lack of immune diversity due to ongoing expansion of activated clones over the lifetime, has been shown to be associated with more severe COVID-19 in adults^24^ and this may add to the evidence base explaining why children usually present with much less severe disease. Also, patients with severe disease have been reported to have a greater number of expanded, low avidity common cold coronavirus specific clones that may contribute to ongoing inflammation^79^.

One limitation of our work is that only a small number of COVID-19 paediatric patients were included due to a very limited number of hospitalised children in the first wave of the pandemic and all of these had severe disease. In view of our interesting findings for neutrophil recruitment in the airways, we note that we isolated PBMCs from blood, thus excluding neutrophils from our analysis. Future studies should address this.

Overall, the significant differences in response to SARS-CoV-2 between children and adults reflect the changes of the immune landscape over developmental time, which in children are dominated by naive and innate responses, whilst those in adults are dominated by adaptive responses, illustrated by expanded lymphocytes clones. These studies may help to pinpoint the triggers of severe disease in adults with a view towards therapeutic intervention.

Our study demonstrates multiple substantially novel insights from paired multi-omics profiling of both airway epithelium and peripheral blood to fill the gap of our understanding of paediatric epithelial and immune development in health and the specific response to COVID-19.

## Data Availability

The data set from our study can be explored interactively through a web portal: https://covid19cellatlas.org. The data object, as a h5ad file, can also be downloaded from the portal page. The processed data will be available for download from Array Express. Counts matrices from bronchial brushings obtained from patients at Northwestern Memorial Hospital, Chicago, are available at GEO, accession number GSE168215.

## Acknowledgments

We acknowledge assistance from Lucy Thorne, Pei Shi Chia, Jana Eliasova, Douglas King, Melanie Heightmann, Michael Marks, Malcolm Avari, Talisa Mistry, Marianne Shaw-Taylor, Ruchira Pereira, Joseph Machta, Julian Lim, Ruth Prendecki, Claire Frauenfelder, James Rudd, Andrew Hall and the Sanger Institute Core Sequencing facility. We thank Richard Jenner and the UCLH/UCL Biomedical Research Centre for the use of their 10X Chromium controller.

We acknowledge funding from the Chan Zuckerberg Initiative (grant 2017-174169) and from Wellcome (WT211276/Z/18/Z and Sanger core grant WT206194). M.Z.N, S.M.J and K.B.M have been funded by the Rosetrees Trust (M944, M35-F2) and from Action Medical Research (GN2911). The project has received funding from the European Union’s Horizon 2020 research and innovation programme under grant agreement No 874656. M.Z.N. acknowledges funding from the Rutherford Fund Fellowship allocated by the MRC, and M.Z.N. and S.M.J. from the UK Regenerative Medicine Platform 2 (MR/5005579/1), the Longfonds BREATH consortium and University College London Hospitals Biomedical Research Centre. M.Y. is funded by The Jikei University School of Medicine. KBW acknowledges funding from University College London, Birkbeck MRC Doctoral Training Programme. S.S. was supported by a Japan Society for the Promotion of Science Overseas Fellowship (310072). R.G.W. was supported by NIH grant U19AI135964 and a GlaxoSmithKline Distinguished Scholar in Respiratory Health grant from the CHEST Foundation. A.V.M. was supported by NIH grant U19AI135964.

NU SCRIPT Study Investigators authors include: A Christine Argento, Catherine A Gao, Alexander V Misharin, GR Scott Budinger, Jane E Dematte, Helen K Donnelly, Nikolay S Markov, Richard G. Wunderink, Sean B Smith, Taylor A Poor, Ziyan Lu.

## Author contributions

M.Z.N. and K.B.M. conceived, set up and directed the study. C.B., E.K., A.W., B.J., A.Sal., A.Sai., H.Y., S.M.J., S.S., P.M., N.S., P.d.C., V.C., J.C., C.C., A.I., M.Z.N. recruited patients, collected samples (where applicable also through bronchoscopies) and clinical metadata.

K.B.W. and M.Y. assisted with sample and meta-data collection, isolated PBMCs and performed single cell isolation of nasal, tracheal and bronchial brushings. K.B.W. and M.Y. performed 10X and CITE-seq, isolated DNA for genotyping. J.A.H. collected samples, performed single cell isolation and 10X (including CITEseq) on post-COVID samples. J.L.B. helped with study set up, CITE-seq and isolated DNA for genotyping. L.M., L.B., L.R. prepared sequencing libraries and conducted the sequencing. E.P. co-ordinated sample shipment and meta-data collection. N.H., R.G.H.L., N.K., C.D.C., E.M., K.P., P.J.P. performed bioinformatic analysis. M.Z.N., K.B.M., K.B.W., M.Y., R.G.H.L., N.H., N.K., C.D.C., E.M., W.S. interpreted the data. K.P. facilitated online data hosting. G.R., M.H. provided help with PBMC annotation. N.J.S., B.J., S.S. provided stored healthy paediatric control nasal tissue blocks. M.C., G.B and M.C carried out experiments to collect and stain post-nasal biopsies. F.J.C.N. and B.G. designed the CITEseq panel and advised on CITEseq experimental design. K.B.M., M.Z.N., K.B.W., M.Y., R.G.H.L., N.H. wrote the manuscript. E.M., S.A.T., B.G., W.S., S.M.J., P.J.P., L.M. edited the manuscript. C.S., C.O., P.d.C., S.M.J., C.B. provided support through ethics and patient recruitment. C.J. and A.K.R. provided support in setting up and training for all CL3 work. Northwestern (bronchial samples): A.C.A., C.A.G., G.R.S.B., J.E.D., R.G.W., S.B.S. and T.A.P. performed bronchoscopies, collection of bronchial brushings and curation of clinical metadata. A.V.M. performed sample processing and analysis. H.K.D obtained informed consent and coordinated sample collection. N.S.M. performed analysis. Z.L. performed sample processing and library construction.

## Code Availability

All data analysis scripts are available on https://github.com/Teichlab/COVID-19paed.

## Supplemental Data

**Extended Data Figure 1: Overview of patient cohort.**

(**a**) Timeline of sample collections from COVID-19 positive (18 adults and 4 paediatric) and post-COVID (4 adults and 2 paediatric) patients enrolled in our study. Sample collections are shown relative to symptom onset and a SARS-CoV-2 positive RT-qPCR test, to which all patients are aligned. (**b**) Overview of samples taken in our post-COVID cohort consisting of patients who were sampled 3 months after recovering from severe COVID-19 (requiring non-invasive or invasive ventilation). (**c**) Ethnicity inferred from scRNA-seq data via principal component analysis of SNP genotypes (red points: our samples; others points: 1000 Genomes Project samples).

**Extended Data Figure 2: Quality control metrics and normalization of airway scRNA-seq dataset.**

UMAP visualisation of annotated airway scRNA-seq dataset (138,688 high quality scRNA-seq transcriptomes) coloured **(a)** by quality control measurements: percentage cell viability based on staining prior to loading cells, UMI count per cell, genes per cell, percentage of mitochondrial gene expression, percentage of hemoglobin expression, percentage of ribosomal expression, percentage of top 50 genes expressed, scrublet score (cell doublet score) and **(b)** by technical factors and metadata: 10X single cell 5’ Chromium kit used (v1.0 or v1.1 NEXT GEM), the hospital patient was recruited from (source), sex, ethnicity, smoking status, Body Mass Index and donor ID. GOSH: Great Ormond Street Hospital, UCLH: University College London Hospital, Royal Free: Royal Free London NHS Foundation Trust, AFR: African, EAS: East Asian, EUR: European, SAS: South Asian, nan: not annotated. **(c)** Violin plots depicting the post-QC distribution of UMI and the percentage of mitochondrial gene expression per cell for each sample.

**Extended Data Figure 3: Expression of viral entry-associated genes in the airways.**

**(a)** Dot plots showing cell type expression of viral entry-associated genes within healthy paediatric airways (n=30), included genes linked to SARS-CoV-2, SARS-CoV, MERS-CoV, Rhinovirus-C and Influenza A infections. The fraction of expressing cells and average expression within each cell type is indicated by dot size and colour, respectively. **(b)** Spearman correlation between average detected level of viral RNA and average expression of entry factors as in (a) across cell types. Dots in red indicate p < 0.05. **(c)** Bar plot comparing fraction of cells expressing *ACE2*, between adult COVID-19 positive, adult post-COVID-19, paediatric COVID-19 positive, paediatric healthy and paediatric post-COVID-19 positive within the airways (nasal and tracheal cells only).

**Extended Data Figure 4:**

**(a)** Expression of “Hillock” cell associated markers (*KRT14*, *KRT6A*, *KRT13* and *KRT24*) was visualised in the epithelial cell UMAP. Highlighted cells belong to the basal, secretory, goblet and squamous cell types, but are a distinct subpopulation. **(b)** Feature importance plot depicting the variance accounted for by each of the clinical and technical factors in our statistical analysis of cell type proportions within our airway scRNA-seq dataset. Factors were tissue type (nasal, tracheal, bronchial), donor ID (patient), date sample was processed (Date_of_process), COVID-19 status (COVID-19 positive, negative or post-COVID-19), patient age (Age_bin), cell viability (cell_viabiity), sex, 10X chromium 5’ single cell sequencing kit version (10X_kit), smoking status, ethnicity and other factors (residual). **(c)** Bar chart showing changes in nasal epithelial cell type proportions observed across age within our paediatric healthy cohort.

**Extended Data Figure 5: Response to interferon.**

Scores of GO term gene signatures for the terms: response to type 1 interferon (GO:0035455 or GO:0034340) and interferon-gamma (GO:0034341) across cell types. Scores were calculated with Scanpy as the average expression of the signature genes subtracted with the average expression of randomly selected genes from bins of corresponding expression values. Scores are shown for both airway **(a)** and PBMC **(b)** data sets. **(c)** Comparison of the gene signature scores between COVID-19 positive adult and COVID-19 positive paediatric groups as well as COVID-19 positive and convalescent adult groups for cells isolated from blood. Results for all cells and monocytes, as the highest responder cell type, are shown. P-values for Wilcoxon rank-sum tests are given on top of each plot.

**Extended Data Figure 6.**

Comparison of “transitioning” epithelial cell populations between our annotated **(a)** upper (nasal and tracheal) and **(b)** lower (bronchi) airway datasets, in addition to **(c)** Chua *et al* 2020 data set^25^. UMAP visualization of deuterosomal (Deu) (*CCNO*, *CDC20B*, *FOXN4*), basal differentiating (Ba-d) (*KRT4*, *KRT7*, *IFT43*) and interferon responsive cell (IRC) (*ISG15*, *IFT1*, *CXCL10*) and inflammatory epithelial transit cell (IETC) (*SAA1*, *S100A8*, *A100A9*) marker gene expression. Ba-d and IRC gene markers used were taken from the Chua *et al* 2020 paper.

**Extended Data Figure 7: Quality control metrics and normalization of PBMC scRNA-seq dataset.**

**(a)** UMAP visualisation of annotated PBMC scRNA-seq dataset (317,854 high quality scRNA-seq transcriptomes) by; number of genes detected (log10), number of proteins detected (log10), gene UMI count per cell (log10), antibodyderived tag UMI count per cell (log10), sex (male, female), sequencing library batch, percentage of mitochondrial gene expression (per read) and sample ID. **(b)** Violin plots depicting the number of gene (top) and antibody derived (bottom) UMIs detected per sample. **(c)** Bar chart showing number of cells detected per sample after filtering (see methods).

**Extended Data Figure 8.**

**(a)** Expanded dotplot (from **Figure 3c**) of matched RNA expression (blue half) and surface protein expression (red half) of cell type markers. The size of the dot is scaled to the percentage of cells that have at least one count of each gene or protein. The colour is scaled to the z-score normalized expression of each gene or protein compared to all other cell types. **(b)** Comparison of our manual cell type PBMC annotation vs an automated annotation performed by Azimuth^62^. **(c)** Trajectory analysis of B naive cells (left) and CD14+ monocytes (right) with age within the blood. Gene expression shown as a z-score and indicated by colour.

**Extended Data Figure 9.**

**(a)** Feature importance plots for cell type proportions of nasal immune cells (left) and in nasal immune cells (middle) and PBMC samples (right) with COVID-19 status calculated separately for adults vs children. **(b)** Dot plot showing marker genes for annotated airway immune cell types, with fraction of expressing cells and average expression within each cell type indicated by dot size and colour, respectively. **(c)** Bar plots showing the average immune cell type proportions collected per patient highlighting the three outliers within the healthy young children (*) presenting B cell hyper infiltration and analysed separately from the rest of the group in **Figure 3f**. Cell types are color coded as shown, with *K* denoting the number of cells per individual.

**Extended Data Figure 10: BCR and TCR clonal expansion (or T and B lymphocyte clonality)**

From our 5’ 10x single cell libraries T cell receptors (TCR) and B cell receptors (BCR) were amplified, sequenced and used to study clonality. T cell subtypes were combined/integrated across all sites (airway and blood) and visualised in an annotated UMAP coloured by cell type **(a)** and TCR chain pairing **(b)**. **(c)** Bar chart showing the (top) total number of (single pair TCR ab chain) T cell clones and (bottom) fraction of TCR with differing clone sizes detected per sample type/location (nasal, PBMC, trachea), where clone size is represented by colour. **(d)** Dandelion plots of T cell clonotypes network by sample ID (left) and sample type/location (right). **(e)** Annotated UMAP of integrated B cell subtypes from across all sites, coloured by receptor subtype (left), cell subtype (middle) and BCR chain pairing (right). **(f)** Bar chart of total number (single pair chain) of B cells clones (top) and fraction of BCRs with differing clone sizes (bottom) detected per sample type/location (nasal, PBMC and tracheal), where clone size is represented by colour. B cell clonotype network by sample ID **(g)** and sample type/location **(h)**.

**Extended Data Table 1: Summary of patient metadata.**

**Extended Data Table 2: TotalSeq-C antibody panel.**

List of 192 TotalSeq-C antibodies (Biolegend, 99814) used for CITE-seq staining of PBMCs; including DNA_ID, Clone, description of target protein and barcode.

**Extended Data Table 3: Viral genomes.**

A complete list of the additional 21 viral genomes used (as FASTA sequences and corresponding GTF entries) during the mapping of single cell data, including their NCBI ID, viral name and source links.

## Methods

### Study Participants and Design

#### The UK cohort

Subjects were included from three large hospital sites in London, United Kingdom, namely Great Ormond Street Hospital NHS Foundation Trust, University College London Hospitals NHS Foundation Trust and Royal Free London NHS Foundation Trust. All healthy paediatric control subjects were obtained from Great Ormond Street Hospital NHS Foundation Trust. Ethical approval was given through the Living Airway Biobank, administered through UCL Great Ormond Street Institute of Child Health (REC reference: 19/NW/0171, IRAS project ID 261511, North West - Liverpool East Research Ethics Committee), REC reference 18/SC/0514 (IRAS project 245471, South Central - Hampshire B Research Ethics Committee) administered through University College London Hospitals NHS Foundation Trust and REC reference 18/EE/0150 (IRAS project ID 236570, East of England - Cambridge Central Research Ethics Committee) administered through Great Ormond Street Hospital NHS Foundation Trust, as well as by the local R&D departments at all hospitals. At daily virtual COVID19 co-ordination meetings suitable patients were chosen from a list of newly diagnosed and admitted patients within the preceding 24 hours (based on a positive nasopharyngeal qRT-PCR test for SARS-CoV-2). Patients with typical clinical and radiological COVID-19 features but with a negative screening test for SARS-CoV-2 were excluded. Other excluding criteria included active haematological malignancy or cancer, known immunodeficiencies, sepsis from any cause and blood transfusion within 4 weeks. We did not include cases of paediatric multisystem inflammatory syndrome (PMIS, named by the Royal College of Paediatrics and Child Health) which is also referred to as multisystem inflammatory syndrome in children (MIS-C) by the World Health Organisation^80^. Maximal severity of COVID-19 was determined retrospectively by determining the presence of symptoms, the need for oxygen supplementation and the level of respiratory support (mild - symptomatic without oxygen requirement or respiratory support, moderate - requiring oxygen without respiratory support, severe - requiring non-invasive or invasive ventilation). Brushings and peripheral blood sampling were performed by trained clinicians prior to inclusion in any pharmacological interventional trials ideally within 48 hours of positive SARS-CoV-2 nasopharyngeal sampling.

#### Chicago Cohort (adult bronchial samples)

Ethical approval for sample collection from patients with severe pneumonia was given by Northwestern Institutional Review Board, study STU00204868 (PI Richard Wunderink). Samples from patients with COVID-19, viral pneumonia and other pneumonia, and non-pneumonia controls were collected from participants enrolled in the Successful Clinical Response in Pneumonia Therapy (SCRIPT) study STU00204868 and admitted to the ICU at Northwestern Memorial Hospital, Chicago. All study participants or their surrogates provided informed consent. Individuals of at least 18 years of age with suspicion of pneumonia based on clinical criteria (including but not limited to fever, radiographic infiltrate and respiratory secretions) were screened for enrolment into the SCRIPT study. Inability to safely perform bronchoalveolar lavage or non-bronchoscopic bronchoalveolar lavage were considered exclusion criteria. In our center, patients with respiratory failure are intubated on the basis of the judgement of bedside clinicians for worsening hypoxaemia, hypercapnia or work of breathing refractory to high-flow oxygen or non-invasive ventilation modes. Extubation occurs on the basis of the judgement of bedside clinicians following a protocol-driven trial of spontaneous breathing in patients demonstrating physiological improvement in their cardiorespiratory status during their period of mechanical ventilation. Bronchial brushings were performed during diagnostic bronchoalveolar lavage procedure and samples were collected from representative sites at the lobar bronchi.

### Sample Collection

#### The UK Cohort

Samples were collected and transferred to a Category Level 3 facility at University College London and processed within 2 hours of sample collection. Nasal, tracheal and bronchial brushings were enzymatically digested to a single cell suspension and processed further immediately. Peripheral blood was centrifuged after adding Ficoll Paque Plus and PBMCs, serum and neutrophils separated, collected and frozen for later processing.

#### Chicago Cohort (adult bronchial samples)

Samples were collected in the ICU at Northwestern Memorial Hospital, transferred to a research laboratory in the Simpson Querrey Biomedical Research Center, Feinberg School of Medicine, Northwestern University, and processed within 1 hour of sample collection in biological safety level 2 facility using biological safety level 3 practices. Upon collection bronchial brushings were stored in Hypothermosol (Stem Cell Technologies, 07935) at 4 °C.

### Nasal and tracheal brushing tissue dissociation

#### The UK Cohort

Nasal brushing was performed on the inferior nasal concha zone with a cytological brush (Scientific Laboratory Supplies, CYT1050). Samples were processed based on protocol from Deprez, Zaragosi *et al*^44^ with minimal modifications. The brushes were immediately placed in a 15 mL sterile falcon tube containing 4mL of transport media on ice. Transport media; αMEM supplemented with 1X penicillin/streptomycin (Gibco; 15070) and 250 ng/ml amphotericin B (Fisher Scientific; 10746254). Once in the Category Level 3 facility, the tube was shaken vigorously to collect cells in suspension. The brushes were then carefully transferred into a new falcon tube containing HBSS and shaken to remove residual cells from the brush. This was repeated until all cells looked like they had been collected from the brush. All falcon tubes were centrifuged at 400 g for 5 min at 4 °C. The cell pellet was collected from each tube and then put in a dissociation buffer consisting of 10 mg/mL protease from Bacillus Licheniformis (Sigma-Aldrich, P5380) and 0.5 mM EDTA in HypoThermosol (Stem Cell Technologies, 07935) for dissociation on ice for 30 min. Every 5 min, cells were gently triturated using a 21 G and 23 G needle. After incubation, protease was inactivated by adding 200 µL of inactivation buffer (HBSS containing 2% BSA). The suspension was centrifuged at 400 g for 5 min at 4 °C and the supernatant was discarded. Cells were resuspended in 1 mL wash buffer (HBSS containing 1% BSA) and centrifuged again. Red blood cell lysis was performed if needed, followed by an additional wash. The single-cell suspension was forced through a 40 µm Flowmi Cell Strainer. Finally, cells were centrifuged and resuspended in 30 µL of resuspension buffer (HBSS containing 0.05% BSA). Using Trypan Blue, total cell counts and viability were assessed. The cell concentration was adjusted for 5000 targeted cell recovery according to the 10X Chromium manual before loading on 10X chip (between 700-1000 cells/µL) and processing immediately for 10X 5’ single cell capture using either the Chromium Single Cell V(D)J Reagent Kits V1.0 (Rev J Guide) or the newer chromium Next GEM Single Cell V(D)J Reagent Kit v1.1 (Rev E Guide).

#### Chicago Cohort (adult bronchial samples)

Samples were processed using the protocol from Deprez, Zaragosi *et al*^44^ with minimal modifications. Specifically, dissociation was performed without EDTA and trituration was performed by pipetting using a regular bore 1000 µL tip every 5 minutes. Dissociation was visually confirmed by inspecting an aliquot of the single cell suspension using phase contrast on an inverted microscope. Cell count was performed using AO/PI reagent on K2 Cellometer (Nexcelom). Approximately 300,000-500,000 cells were obtained per brush with viability of 97% and above. Cells were captured on a 10X Chromium Single Cell Controller using Chromium Single Cell V(D)J Reagent Kits V1.0 (Rev J Guide).

### PBMC isolation from peripheral blood

Peripheral blood was collected in EDTA immediately after the nasal brushing procedure. The blood was diluted with 5 mL of PBS containing 2 mM EDTA (Invitrogen, 1555785-038). 10 to 20 mL of diluted blood was carefully layered onto 15 mL of Ficoll-Paque Plus (GE healthcare, 17144002). If the sample volume was less than 5 mL, blood was diluted with an equal volume of PBS-EDTA and layered onto 3 mL Ficoll. The sample was centrifuged at 800 g for 20 min at room temperature. The plasma layer was carefully removed and the peripheral blood mononuclear cell (PBMC) layer was collected using a sterile Pasteur pipette. The PBMC layer was washed with 3 volumes of PBS containing EDTA by centrifugation at 500 g for 10 min. The pellet was suspended in PBS-EDTA and centrifuged again at 300 g for 5 min. The PBMC pellet was collected followed by both cell number and viability being assessed using Trypan Blue. Cell freezing medium (90% FBS, 10% DMSO) was added dropwise to PBMCs slowly on ice and then the mixture was cryopreserved at -80 °C until further full sample processing.

### CITE-Seq staining for single-cell proteogenomics

Frozen PBMC samples were thawed quickly at 37 °C in a water bath. 20-30 mL of warm RPMI1640 medium containing 10% FBS was added slowly to the cells before centrifuging at 300 g for 5 min. This was followed by a wash in 5 mL RPMI1640-FBS. The PBMC pellet was collected, and cell number and viability were determined using Trypan Blue. PBMCs from four different donors were then pooled together at equal numbers - 1.25x10^5^ PBMCs from each donor were combined with the other PBMCs to make up 5.0x10^5^ cells in total. The remaining cells were used for DNA extraction (Qiagen, 69504). The pooled PBMCs were resuspended in 25 µL of cell staining buffer (Biolegend, 420201) and blocked via a 10 minute incubation on ice with 2.5 µL Human TruStain FcX block ( Biolegend, 422301). The PBMCs pool was then stained with TotalSeq-C antibodies (Biolegend, 99814) according to manufacturer’s instructions. The full list of TotalSeq-C antibodies can be found in **Extended Data Table 2**. After incubating with 0.5 vial of TotalSeq-C for 30 min at 4 °C, PBMCs were washed three times by centrifugation at 500 g for 5 min at 4 °C. PBMCs were counted again and processed immediately for 10X 5’ single cell capture (Chromium Next GEM Single Cell V(D)J Reagent Kit v1.1 with Feature Barcoding technology for cell Surface Protein-Rev D protocol). Two lanes of 25,000 cells were loaded per pool on a 10X chip.

### Library Generation and Sequencing

Either Chromium Single Cell 5’ V(D)J Reagent Kit (V1.0 chemistry) or Chromium Next GEM Single Cell 5’ V(D)J Reagent Kit (V1.1 chemistry) was used for scRNAseq library construction for all airway samples, with Chromium Next GEM Single Cell V(D)J Reagent Kit v1.1 with Feature Barcoding technology for cell surface proteins used for PBMCs. Gene expression libraries (GEX) and V(D)J libraries were prepared according to the manufacturer’s protocol (10X Genomics) using individual Chromium i7 Sample Indices. The cell surface protein libraries were created according to the manufacturer’s protocol with slight modification that included doubling the SI primer amount per reaction and reducing the number of amplification cycles to 7 during the index PCR to avoid the daisy chains effect. GEX, V(D)J and cell surface protein indexed libraries were pooled in 1:0.1:0.4 ratio respectively and sequenced on a NovaSeq 6000 S4 Flowcell (paired-end (PE), 150-bp reads) aiming for a minimum of 50,000 PE reads per cell for GEX libraries and 5,000 PE reads per cell for V(D)J and cell surface protein libraries.

### Single Cell RNA-seq Computational Pipelines, Processing and Analysis

The single cell data was mapped to a GRCh38 ENSEMBL 93 derived reference, with an additional 21 viral genomes (featuring SARS-CoV2) included as additional FASTA sequences and corresponding GTF entries. A complete list of the included viruses, along with their respective NCBI IDs and source links, can be found in **Extended Data Table 3**. Antibody-derived tag counts (ADT) and gene expression counts in CITE-seq data were jointly quantified using Cellranger 3.0.2. The alignment, quantification and preliminary cell calling of airway samples were carried out via the STARsolo functionality of STAR 2.7.3a, with the cell calling subsequently refined with Cell Ranger 3.0.2’s version of EmptyDrops^81^. This algorithm has been made available as emptydrops on PyPi. Initial doublets were called on a per-sample basis by computing Scrublet^82^ scores for each cell, propagating them through an over-clustered manifold by replacing individual scores with per-cluster medians, and identifying statistically significant values from the resulting distribution, replicating the approach of Pijuan-Sala *et al*, 2019^83^ and Popescu *et al*, 2019^84^. The clustering was performed with the Leiden^61^ algorithm on a KNN graph of a PCA space derived from a log(CPM/100 + 1) representation of highly variable genes, following SCANPY protocol^85^, and overclustering was achieved by performing an additional clustering of each resulting cluster. The primary clustering also served as input for ambient RNA removal via SoupX^86^.

### Confocal microscopy method

Nasal epithelial biopsies were placed in Antigenfix (Microm Microtech) for 1-2 h at 4°C, then 30% sucrose in PBS for 12-24 h at 4°C, before cryopreservation in OCT (Cell Path). 30µm sections were permeabilised and blocked in PBS containing 0.3% Triton (Sigma), 1% normal goat serum, 1% normal donkey serum, and 1% BSA (R&D) for 1-2 h at room temperature (RT). Samples were stained with a 1 in 50 dilution of anti-human S100A9 conjugated to FITC (clone: MRP 1H9, Biolegend) and a 1 in 50 dilution of anti-human EpCam conjugated to APC (clone: 9C4, Biolegend) in blocking buffer overnight and washed for 3 x 10 minutes in PBS before mounting with Fluoromount-G containing DAPI (Invitrogen). Images were acquired using a Leica SP8 confocal microscope. Raw imaging data were processed using Imaris (Bitplane).

### Airway single cell RNA-seq data processing

#### Quality control and normalization

To account for large quality variance across different samples, quality control was done for each sample separately. QC thresholds were automatically established by fitting a 10- component Gaussian mixture model to log-transformed UMI count per cell and to percentage of mitochondrial gene expression and finding the lower or higher bounds where probability density falls under 0.05. For three samples (NP32-NB, NP39-NB and AP10-NB) for which the automatic procedure failed to find reasonable thresholds, the distribution of quality metrics were examined and thresholds were determined manually. We also excluded genes expressed in fewer than 3 cells. Expression values were then normalised to a sum of 1e4 per cell and log transformed with an added pseudo-counting of 1.

#### Developmental trajectory inference

RNA velocity analysis was performed to infer developmental trajectory for the major epithelial cell types (excluding melanocytes, ionocytes, brush cells and neuroendocrine cells). Spliced and unspliced UMI counts were generated via the STARsolo functionality of STAR 2.7.3a. scvelo was used to fit a dynamical model as previously described^87^, based on top 2,000 highly variable genes with at least 20 UMI for both spliced and unspliced transcripts across all cells.

### CITE-seq data processing

#### Demultiplexing and doublet removal of PBMC samples

For pooled donor CITE-seq samples, the donor ID of each cell was determined by genotype-based demultiplexing using souporcell version 2 ^88^. Souporcell analyses were performed with ‘skip_remap’ enabled and a set of known donor genotypes given under the ‘common_variants’ parameter. The donor ID of each souporcell genotype cluster was annotated by comparing each souporcell genotype to the set of known genotypes. Droplets that contained more than one genotype according to souporcell were flagged as ‘ground-truth’ doublets for heterotypic doublet identification. Ground-truth doublets were used by DoubletFinder 2.0.3 ^89^ to empirically determine an optimal ‘pK’ value for doublet detection. DoubletFinder analysis was performed on each sample separately using 10 principal components, a ‘pN’ value of 0.25, and the ‘nExp’ parameter estimated from the fraction of ground-truth doublets and the number of pooled donors.

#### CITE-seq background and ambient RNA subtraction

Background and non-specific staining by the antibodies used in CITE-seq was estimated using SoupX version 1.4.8^86^, which models background signal on near-empty droplets. The ‘soupQuantile’ and ‘tfidfMin’ parameters were set to 0.25 and 0.2, respectively, and lowered by decrements of 0.05 until the contamination fraction was calculated using the ‘autoEstCont’ function. Gene expression data was also corrected with SoupX to remove cell free mRNA contamination using default SoupX parameters.

#### CITE-seq quality control and normalization

CITE-seq data was filtered by removing droplets with fewer than 200 genes expressed or with more than 10% of the counts originating from mitochondrial genes. Gene expression data was normalized with a log + 1 transformation (log1p), and 2000 hyper variable genes were selected with the vst algorithm in Seurat version 3.9.9.9024^90^. Antibody derived tag counts were normalized with the centered log-ratio (CLR) transformation.

#### Integrated embedding and clustering of CITE-seq data

Principal component analysis was run separately on gene expression and antibody derived tag count data, followed by batch correction using harmony^91^ on the sequencing library identifier. Nearest neighbor graphs and uniform manifold approximation and projections (UMAP) were generated based on the first 30 harmony-adjusted principal components. The first 30 harmony-adjusted principal components of both gene expression and antibody derived tag count data were used to compute a weighted nearest neighbor (WNN) graph [https://doi.org/10.1101/2020.10.12.335331] with Seurat and embedded using UMAP. Cells were clustered with the Leiden algorithm using the igraph R package, with a resolution of 4. After initial clustering of all PBMCs, subsets of all T and NK cells, all B and plasma cells, and all monocytes and DCs were reclustered after hypervariable gene selection within each subset. Cells in WNN-based clusters with less than 100 members were reassigned based on the closest multimodal neighbour.

#### Comparison PBMCs and Azimuth

The manual blood cell-type annotation was validated using the Azimuth tool (http://azimuth.satijalab.org/app/azimuth). A randomly sampled subset of 100,000 PBMCs were uploaded to predict their cell type identity. Exhausted B, switched B, non-switched B, complement expressing monocytes, NKT and plasma cells in our annotation were not present in the Azimuth annotation, while intermediate B, ASDC and double negative T cells were not present in our annotation.

#### Differential expression analysis in naïve T cells, B cells and monocytes

Two subsets of all naïve CD4 and CD8 T cells from healthy paediatric donors were reclustered after hypervariable gene selection within each subset to generate the UMAP in Figure 3d. Differential gene expression across age within these subsets was tested by fitting a gamma-poisson generalized linear model on log_2_ transformed age and by creating pseudo-bulks of each donor with the glmGamPoi package^91, 92^ [https://doi.org/10.1093/bioinformatics/btaa1009].

#### Differential expression analysis in airway data

In addition to the differential expression analysis correcting for various metadata that was performed on the whole airway and PBMC data sets as described below, results shown for subsets of the data were obtained with a simpler method. After subsetting cell types and/or age groups, a Wilcoxon rank-sum test (implemented in Scanpy^85^) was performed to compare gene groups. The sets of differentially expressed genes were further analysed with the g:Profiler toolkit^93^ (g:Profiler version *e102_eg49_p15_7a9b4d6*, database updated on 15/12/2020) for functional enrichment analysis.

#### Correlation analysis on circulating versus resident immune cells

To harmonize the cell type annotation in the airway samples with the manual annotation of cell in PBMC samples, we integrated the airway immune cells with the PBMC data by IntegrateData function in Seurat on the top 30 principal components. Immune cell types that were unique to the airway samples were (activated) macrophages, neutrophils, follicular DCs and mast cells, and were therefore manually annotated. Airway CD8 T cells were annotated as CD8 naïve or effector cells because further CD8 effector subtypes were less pronounced in airway samples. The relative immune cell type proportions in matched nasal and blood samples were correlated using the Spearman rank-order method, where we only tested the correlation of cell types that were present in at least half of the nasal samples in each comparison.

#### Inference of ethnicity from single cell RNA-seq data

The latest biallelic SNP genotype data (GRCh38) was obtained from 1000 Genomes Project (see URL: ftp://ftp.1000genomes.ebi.ac.uk/vol1/ftp/data_collections/1000_genomes_project/release/201 81203_biallelic_SNV/). Allele specific counts of RNA-seq reads at the SNP location in 1000 Genomes Project data were generated for each airway sample. Because the read coverage from the scRNA-seq data was strongly enriched around the 5’ end of a gene, SNP loci covered at least 20 reads for more than 90% of samples that were used (19,733 genome-wide SNP loci in total). The SNP genotype from allele specific expression was determined as a maximum posterior genotype after fitting a beta-binomial mixture distribution with underlying probabilities of 0.01, 0.5 and 0.99 for reference homozygote, heterozygote and alternative homozygote, respectively. The overdispersion parameter of the beta-binomial distribution was estimated for each sample independently shared across all SNPs. The genotype data from 1000 Genomes samples were combined with the genotype data for our samples, and principal component analysis was performed on the scaled genotype data (mean 0 and standard deviation equal to 1 for each SNP locus). The ethnicity of each sample was determined by the Mahalanobis distance to the four major ethnic groups in the 1000 Genomes Project (African, East Asian, European and South Asian). The first three principal components were used to compute the cluster centre and the covariance matrix for each ethnic group.

#### Cell type composition analysis

The number of cells for each sample and cell type combination was modelled with a generalised linear mixed model with a Poisson outcome. The five clinical factors (age, sex, ethnicity, tissue and COVID-19 status) and the six technical factors (donor, date of sample processing, 10X kit, technician, sequencing batch and sample) were fitted as random effects to overcome the colinearity among the factors. The effect of each clinical/technical factor on cell type composition was estimated by the interaction term with the cell type. The ‘glmer’ function in the lme4 package implemented on R was used to fit the model. The standard error of variance parameter for each factor was estimated using the numDeriv package. The conditional distribution of the fold change estimate of a level of each factor was obtained by the ‘ranef’ function in the lme4 package. The statistical significance of the fold change estimate was measured by the local true sign rate (LTSR) which is an area under the curve of the posterior distribution of a log fold change over the positive domain when the point estimate is greater than 0 (over the negative domain for the point estimate less than 0).

#### Differential expression analysis using metadata

We performed differential gene expression analysis for both airway and PBMC data. We use the 7 clinical (Donor, Age group, Sex, Ethnicity, Tissue, Smoking status and COVID-19 status) and the 4 technical factors (Batch, 10X kit version, the number of expressed genes and the number of mapped fragments) to adjust confounding effects. For PBMC data, the tissue and 10X kit are identical across samples and not included in the model. We utilised the linear mixed model proposed in Young *et al*^94^ to adjust for the 11 confounding factor effects and the effect of cell type as a random effect in differential expression analysis. We fit the model on a gene-by-gene basis using the estimated variance parameters to test each factor k explaining a significant amount of transcription variation. If the focal factor *k* is a categorical variable with *L* levels (e.g., COVID-19 status with 3 levels), we partitioned the levels into one of two groups. There are 2*L*-1 contrasts which were tested against the null model (removing the focal factor *k* in the model) to compute Bayes factors. Then, those Bayes factors were used for fitting a finite mixture model to compute the posterior probability as well as the local true sign rate (ltsr) (See Supplementary Note of Young *et al*^94^ Section 1.3 for more details). We used g:Profiler 2 implemented in R (version 2.0.1.5) to identify which pathways are enriched for differentially expressed genes for each contrast. We used genes whose ltsr is greater than 0.5 to perform the analysis (both upregulated and downregulated genes separately).

#### Single-cell VDJ-sequencing data analysis

TCR and BCR sequencing data was processed using the Cellranger software and downstream analysis was performed using the scirpy package (version 0.6.1)^95^. Briefly, we integrated TCR and BCR data with gene expression from T cell and B cell subsets, respectively. After categorizing cells based on the detection of productive antigen receptor chains, we selected cells with a single pair of productive chains for further analysis. Clonotypes were defined at the nucleotide level and considering both receptor chains.

## References

1. COVID-19 Cell Atlas. https://www.covid19cellatlas.org/.

2. World Health Organization. Report of the WHO-China Joint Mission on Coronavirus Disease 2019 (COVID-19). https://www.who.int/docs/default-source/coronaviruse/who-china-joint-mission-on-covid-19-final-report.pdf (2020, February 28).

3. Swann, O. V. et al. Clinical characteristics of children and young people admitted to hospital with covid-19 in United Kingdom: prospective multicentre observational cohort study. BMJ 370, m3249 (2020).

4. Xu, Y. et al. Characteristics of pediatric SARS-CoV-2 infection and potential evidence for persistent fecal viral shedding. Nat. Med. 26, 502–505 (2020).

5. Lu, X. et al. SARS-CoV-2 Infection in Children. N. Engl. J. Med. 382, 1663–1665 (2020).

6. Castagnoli, R. et al. Severe Acute Respiratory Syndrome Coronavirus 2 (SARS-CoV-2) Infection in Children and Adolescents: A Systematic Review. JAMA Pediatr. 174, 882– 889 (2020).

7. Rodriguez-Morales, A. J. et al. Clinical, laboratory and imaging features of COVID-19: A systematic review and meta-analysis. Travel Med. Infect. Dis. 34, 101623 (2020).

8. Website. Huang C Wang Y Li X et al. Clinical features of patients infected with 2019 novel coronavirus in Wuhan. China Lancet. 2020; 395: 497-506https://doi.org/10.1016/S0140-6736(20)30183-5.

9. Hoang, A. et al. COVID-19 in 7780 pediatric patients: A systematic review. EClinicalMedicine 24, 100433 (2020).

10. Sisk, B., Cull, W., Harris, J. M., Rothenburger, A. & Olson, L. National Trends of Cases of COVID-19 in Children Based on US State Health Department Data. Pediatrics 146, (2020).

11. Hoffmann, M. et al. SARS-CoV-2 Cell Entry Depends on ACE2 and TMPRSS2 and Is Blocked by a Clinically Proven Protease Inhibitor. Cell 181, 271–280.e8 (2020).

12. Pang, L. et al. Influence of aging on deterioration of patients with COVID-19. Aging 12, 26248–26262 (2020).

13. Muus, C. et al. Integrated analyses of single-cell atlases reveal age, gender, and smoking status associations with cell type-specific expression of mediators of SARS-CoV-2 viral entry and highlights inflammatory programs in putative target cells. bioRxiv 2020.04.19.049254 (2020) doi:10.1101/2020.04.19.049254.

14. Sungnak, W. et al. SARS-CoV-2 entry factors are highly expressed in nasal epithelial cells together with innate immune genes. Nat. Med. (2020) doi:10.1038/s41591-020-0868-6.

15. Bunyavanich, S., Do, A. & Vicencio, A. Nasal Gene Expression of Angiotensin-Converting Enzyme 2 in Children and Adults. JAMA 323, 2427–2429 (2020).

16. Saheb Sharif-Askari, N. et al. Airways Expression of SARS-CoV-2 Receptor, ACE2, and TMPRSS2 Is Lower in Children Than Adults and Increases with Smoking and COPD. Mol Ther Methods Clin Dev 18, 1–6 (2020).

17. Koch, C. M. et al. Immune response to SARS-CoV-2 in the nasal mucosa in children and adults. bioRxiv (2021) doi:10.1101/2021.01.26.21250269.

18. Berlin, D. A., Gulick, R. M. & Martinez, F. J. Severe Covid-19. N. Engl. J. Med. 383, 2451–2460 (2020).

19. Guo, C. et al. Single-cell analysis of two severe COVID-19 patients reveals a monocyte-associated and tocilizumab-responding cytokine storm. Nat. Commun. 11, 3924 (2020).

20. Liao, M. et al. Single-cell landscape of bronchoalveolar immune cells in patients with COVID-19. Nat. Med. 26, 842–844 (2020).

21. Zhou, Y. et al. Pathogenic T-cells and inflammatory monocytes incite inflammatory storms in severe COVID-19 patients. Natl Sci Rev 7, 998–1002 (2020).

22. Wilk, A. J. et al. A single-cell atlas of the peripheral immune response in patients with severe COVID-19. Nat. Med. 26, 1070–1076 (2020).

23. Zhang, J.-Y. et al. Single-cell landscape of immunological responses in patients with COVID-19. Nat. Immunol. 21, 1107–1118 (2020).

24. Stephenson, E. et al. The cellular immune response to COVID-19 deciphered by single cell multi-omics across three UK centres. bioRxiv (2021) doi:10.1101/2021.01.13.21249725.

25. Chua, R. L. et al. COVID-19 severity correlates with airway epithelium-immune cell interactions identified by single-cell analysis. Nat. Biotechnol. (2020) doi:10.1038/s41587-020-0602-4.

26. Pierce, C. A. et al. Immune responses to SARS-CoV-2 infection in hospitalized pediatric and adult patients. Sci. Transl. Med. (2020) doi:10.1126/scitranslmed.abd5487.

27. Weisberg, S. P. et al. Distinct antibody responses to SARS-CoV-2 in children and adults across the COVID-19 clinical spectrum. Nat. Immunol. 22, 25–31 (2021).

28. Lee, A. H. et al. Dynamic molecular changes during the first week of human life follow a robust developmental trajectory. Nat. Commun. 10, 1092 (2019).

29. Olin, A. et al. Stereotypic Immune System Development in Newborn Children. Cell 174, 1277–1292.e14 (2018).

30. Lambert, L. & Culley, F. J. Innate Immunity to Respiratory Infection in Early Life. Frontiers in Immunology vol. 8 (2017).

31. Ygberg, S. & Nilsson, A. The developing immune system - from foetus to toddler. Acta Paediatr. 101, 120–127 (2012).

32. Zierk, J. et al. Next-generation reference intervals for pediatric hematology. Clin. Chem. Lab. Med. 57, 1595–1607 (2019).

33. Syrimi, E. et al. Age related defects in NK cell immunity revealed by deep immune profiling of pediatric cancer patients. doi:10.1101/2020.03.09.983288.

34. Bellussi, L., Cambi, J. & Passali, D. Functional maturation of nasal mucosa: role of secretory immunoglobulin A (SIgA). Multidiscip. Respir. Med. 8, 46 (2013).

35. Grant, R. A. et al. Circuits between infected macrophages and T cells in SARS-CoV-2 pneumonia. Nature 590, 635–641 (2021).

36. Cantuti-Castelvetri, L. et al. Neuropilin-1 facilitates SARS-CoV-2 cell entry and infectivity. Science 370, 856–860 (2020).

37. Daly, J. L. et al. Neuropilin-1 is a host factor for SARS-CoV-2 infection. Science 370, 861–865 (2020).

38. Wang, K. et al. SARS-CoV-2 invades host cells via a novel route: CD147-spike protein. Cold Spring Harbor Laboratory 2020.03.14.988345 (2020) doi:10.1101/2020.03.14.988345.

39. Tang, X. et al. Transferrin receptor is another receptor for SARS-CoV-2 entry. Cold Spring Harbor Laboratory 2020.10.23.350348 (2020) doi:10.1101/2020.10.23.350348.

40. Ziegler, C. G. K. et al. SARS-CoV-2 Receptor ACE2 Is an Interferon-Stimulated Gene in Human Airway Epithelial Cells and Is Detected in Specific Cell Subsets across Tissues. Cell 181, 1016–1035.e19 (2020).

41. Wang, W. et al. Detection of SARS-CoV-2 in Different Types of Clinical Specimens. JAMA 323, 1843–1844 (2020).

42. Yu, F. et al. Quantitative Detection and Viral Load Analysis of SARS-CoV-2 in Infected Patients. Clin. Infect. Dis. 71, 793–798 (2020).

43. Vieira Braga, F. A. et al. A cellular census of human lungs identifies novel cell states in health and in asthma. Nat. Med. 25, 1153–1163 (2019).

44. Deprez, M. et al. A Single-cell Atlas of the Human Healthy Airways. Am. J. Respir. Crit. Care Med. (2020).

45. Travaglini, K. J. et al. A molecular cell atlas of the human lung from single-cell RNA sequencing. Nature 587, 619–625 (2020).

46. Zak, F. G. & Lawson, W. The presence of melanocytes in the nasal cavity. Ann. Otol. Rhinol. Laryngol. 83, 515–519 (1974).

47. Ewing, E. Malignant Melanoma Arising in Association with Sinonasal Melanosis: A Case Report and Review of the Literature. International Journal of Pathology and Clinical Research vol. 3 (2017).

48. Tata, P. R. & Rajagopal, J. Plasticity in the lung: making and breaking cell identity. Development 144, 755–766 (2017).

49. Montoro, D. T. et al. A revised airway epithelial hierarchy includes CFTR-expressing ionocytes. Nature 560, 319–324 (2018).

50. Ziegler, C. G. K. et al. Impaired local intrinsic immunity to SARS-CoV-2 infection in severe COVID-19. bIoRxiv 2021.02.20.431155 (2021) doi:10.1101/2021.02.20.431155.

51. Zhu, N. et al. Morphogenesis and cytopathic effect of SARS-CoV-2 infection in human airway epithelial cells. Nat. Commun. 11, 3910 (2020).

52. Fang, Y. et al. Distinct stem/progenitor cells proliferate to regenerate the trachea, intrapulmonary airways and alveoli in COVID-19 patients. Cell Res. 30, 705–707 (2020).

53. Ruiz García, S. et al. Novel dynamics of human mucociliary differentiation revealed by single-cell RNA sequencing of nasal epithelial cultures. Development 146, (2019).

54. Lazear, H. M., Schoggins, J. W. & Diamond, M. S. Shared and Distinct Functions of Type I and Type III Interferons. Immunity 50, 907–923 (2019).

55. Blanco-Melo, D. et al. Imbalanced Host Response to SARS-CoV-2 Drives Development of COVID-19. Cell 181, 1036–1045.e9 (2020).

56. Chen, G. et al. Clinical and immunological features of severe and moderate coronavirus disease 2019. J. Clin. Invest. 130, 2620–2629 (2020).

57. Sack, G. H., Jr. Serum amyloid A - a review. Mol. Med. 24, 46 (2018).

58. Silvin, A. et al. Elevated Calprotectin and Abnormal Myeloid Cell Subsets Discriminate Severe from Mild COVID-19. Cell 182, 1401–1418.e18 (2020).

59. Aschenbrenner, A. C. et al. Disease severity-specific neutrophil signatures in blood transcriptomes stratify COVID-19 patients. Genome Med. 13, 7 (2021).

60. Hao, Y. et al. Integrated analysis of multimodal single-cell data. bioRxiv 2020.10.12.335331 (2020) doi:10.1101/2020.10.12.335331.

61. Traag, V. A., Waltman, L. & van Eck, N. J. From Louvain to Leiden: guaranteeing well-connected communities. Sci. Rep. 9, 5233 (2019).

62. LSatija Lab. https://satijalab.org/azimuth/.

63. Schmidleithner, L. et al. Enzymatic Activity of HPGD in Treg Cells Suppresses Tconv Cells to Maintain Adipose Tissue Homeostasis and Prevent Metabolic Dysfunction. Immunity 50, 1232–1248.e14 (2019).

64. Aliahmad, P., Kadavallore, A., de la Torre, B., Kappes, D. & Kaye, J. TOX is required for development of the CD4 T cell lineage gene program. J. Immunol. 187, 5931–5940 (2011).

65. Cerutti, A., Cols, M. & Puga, I. Marginal zone B cells: virtues of innate-like antibody-producing lymphocytes. Nat. Rev. Immunol. 13, 118–132 (2013).

66. Timens, W., Boes, A., Rozeboom-Uiterwijk, T. & Poppema, S. Immaturity of the human splenic marginal zone in infancy. Possible contribution to the deficient infant immune response. J. Immunol. 143, 3200–3206 (1989).

67. Xia, W. et al. Clinical and CT features in pediatric patients with COVID-19 infection: Different points from adults. Pediatric Pulmonology vol. 55 1169–1174 (2020).

68. Cui, X. et al. Children with coronavirus disease 2019: A review of demographic, clinical, laboratory, and imaging features in pediatric patients. J. Med. Virol. 92, 1501– 1510 (2020).

69. Li, H., Chen, K., Liu, M., Xu, H. & Xu, Q. The profile of peripheral blood lymphocyte subsets and serum cytokines in children with 2019 novel coronavirus pneumonia. J. Infect. 81, 115–120 (2020).

70. Schulte-Schrepping, J. et al. Severe COVID-19 Is Marked by a Dysregulated Myeloid Cell Compartment. Cell 182, 1419–1440.e23 (2020).

71. Gibbs, A. et al. MAIT cells reside in the female genital mucosa and are biased towards IL-17 and IL-22 production in response to bacterial stimulation. Mucosal Immunol. 10, 35–45 (2017).

72. Sobkowiak, M. J. et al. Tissue-resident MAIT cell populations in human oral mucosa exhibit an activated profile and produce IL-17. Eur. J. Immunol. 49, 133–143 (2019).

73. 73. Parrot, T. et al. MAIT cell activation and dynamics associated with COVID-19 disease severity. *Sci Immunol* 5, (2020).

74. Jouan, Y. et al. Phenotypical and functional alteration of unconventional T cells in severe COVID-19 patients. J. Exp. Med. 217, (2020).

75. Peng, Y. et al. Broad and strong memory CD4 and CD8 T cells induced by SARS-CoV-2 in UK convalescent individuals following COVID-19. Nat. Immunol. 21, 1336–1345 (2020).

76. Yu, J. C. et al. Innate Immunity of Neonates and Infants. Frontiers in Immunology vol. 9 (2018).

77. Li, S. et al. SARS-CoV-2 triggers inflammatory responses and cell death through caspase-8 activation. Signal Transduct Target Ther 5, 235 (2020).

78. Upton, J. W., Shubina, M. & Balachandran, S. RIPK3-driven cell death during virus infections. Immunol. Rev. 277, 90–101 (2017).

79. Bacher, P. et al. Low-Avidity CD4 T Cell Responses to SARS-CoV-2 in Unexposed Individuals and Humans with Severe COVID-19. Immunity 53, 1258–1271.e5 (2020).

80. Lee, P. Y. et al. Distinct clinical and immunological features of SARS-CoV-2-induced multisystem inflammatory syndrome in children. J. Clin. Invest. 130, 5942–5950 (2020).

81. Lun, A. T. L. et al. EmptyDrops: distinguishing cells from empty droplets in droplet-based single-cell RNA sequencing data. Genome Biol. 20, 63 (2019).

82. Wolock, S. L., Lopez, R. & Klein, A. M. Scrublet: Computational Identification of Cell Doublets in Single-Cell Transcriptomic Data. Cell Syst 8, 281–291.e9 (2019).

83. Pijuan-Sala, B. et al. A single-cell molecular map of mouse gastrulation and early organogenesis. Nature 566, 490–495 (2019).

84. Popescu, D.-M. et al. Decoding human fetal liver haematopoiesis. Nature 574, 365–371 (2019).

85. Wolf, F. A., Angerer, P. & Theis, F. J. SCANPY: large-scale single-cell gene expression data analysis. Genome Biol. 19, 15 (2018).

86. Young, M. D. & Behjati, S. SoupX removes ambient RNA contamination from droplet-based single-cell RNA sequencing data. Gigascience 9, (2020).

87. Bergen, V., Lange, M., Peidli, S., Wolf, F. A. & Theis, F. J. Generalizing RNA velocity to transient cell states through dynamical modeling. Nat. Biotechnol. 38, 1408–1414 (2020).

88. Heaton, H. et al. Souporcell: robust clustering of single-cell RNA-seq data by genotype without reference genotypes. Nat. Methods 17, 615–620 (2020).

89. McGinnis, C. S., Murrow, L. M. & Gartner, Z. J. DoubletFinder: Doublet Detection in Single-Cell RNA Sequencing Data Using Artificial Nearest Neighbors. Cell Syst 8, 329– 337.e4 (2019).

90. 90. Stuart, T. et al. Comprehensive Integration of Single-Cell Data. *Cell* vol. 177 1888–1902.e21 (2019).

91. Korsunsky, I. et al. Fast, sensitive and accurate integration of single-cell data with Harmony. Nat. Methods 16, 1289–1296 (2019).

92. Ahlmann-Eltze, C. & Huber, W. glmGamPoi: Fitting Gamma-Poisson Generalized Linear Models on Single Cell Count Data. Bioinformatics (2020) doi:10.1093/bioinformatics/btaa1009.

93. Reimand, J. et al. g:Profiler-a web server for functional interpretation of gene lists (2016 update). Nucleic Acids Res. 44, W83–9 (2016).

94. Young, A. et al. A map of transcriptional heterogeneity and regulatory variation in human microglia. bioRxiv 2019.12.20.874099 (2019) doi:10.1101/2019.12.20.874099.

95. Sturm, G. et al. Scirpy: a Scanpy extension for analyzing single-cell T-cell receptor-sequencing data. Bioinformatics (2020) doi.org/10.1093/bioinformatics/btaa611

