## Extended Data for "The local and systemic response to SARS-CoV-2 infection in children and adults"

### Extended Data Figure 1

a

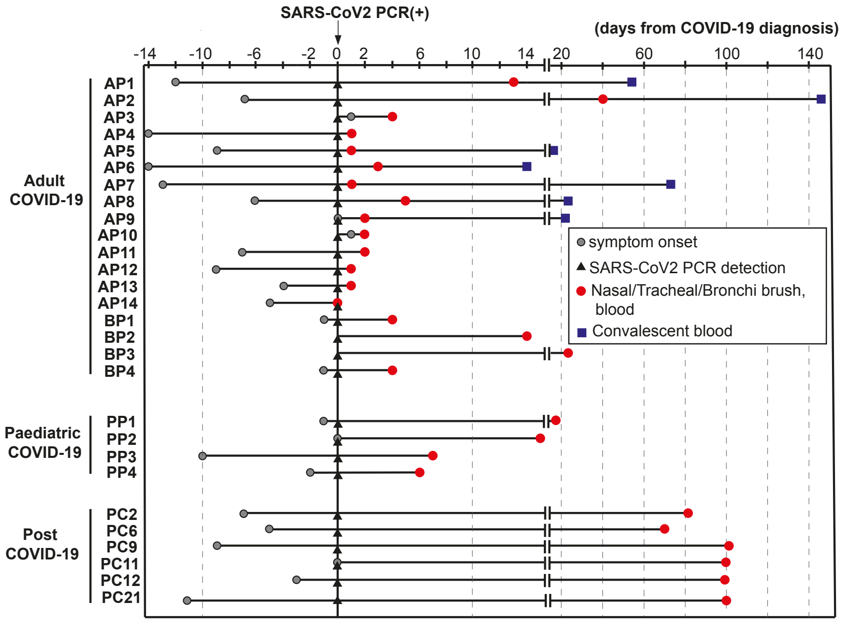


b

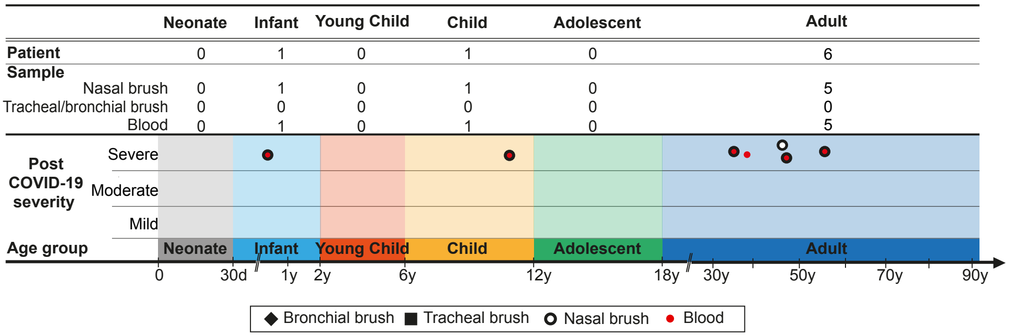


c

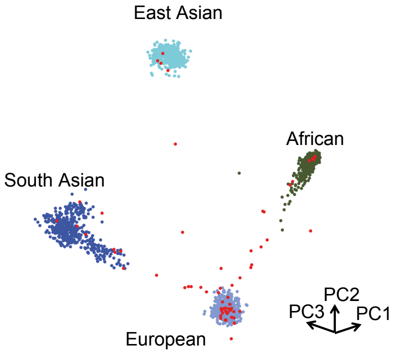

#### Extended Data Figure 2

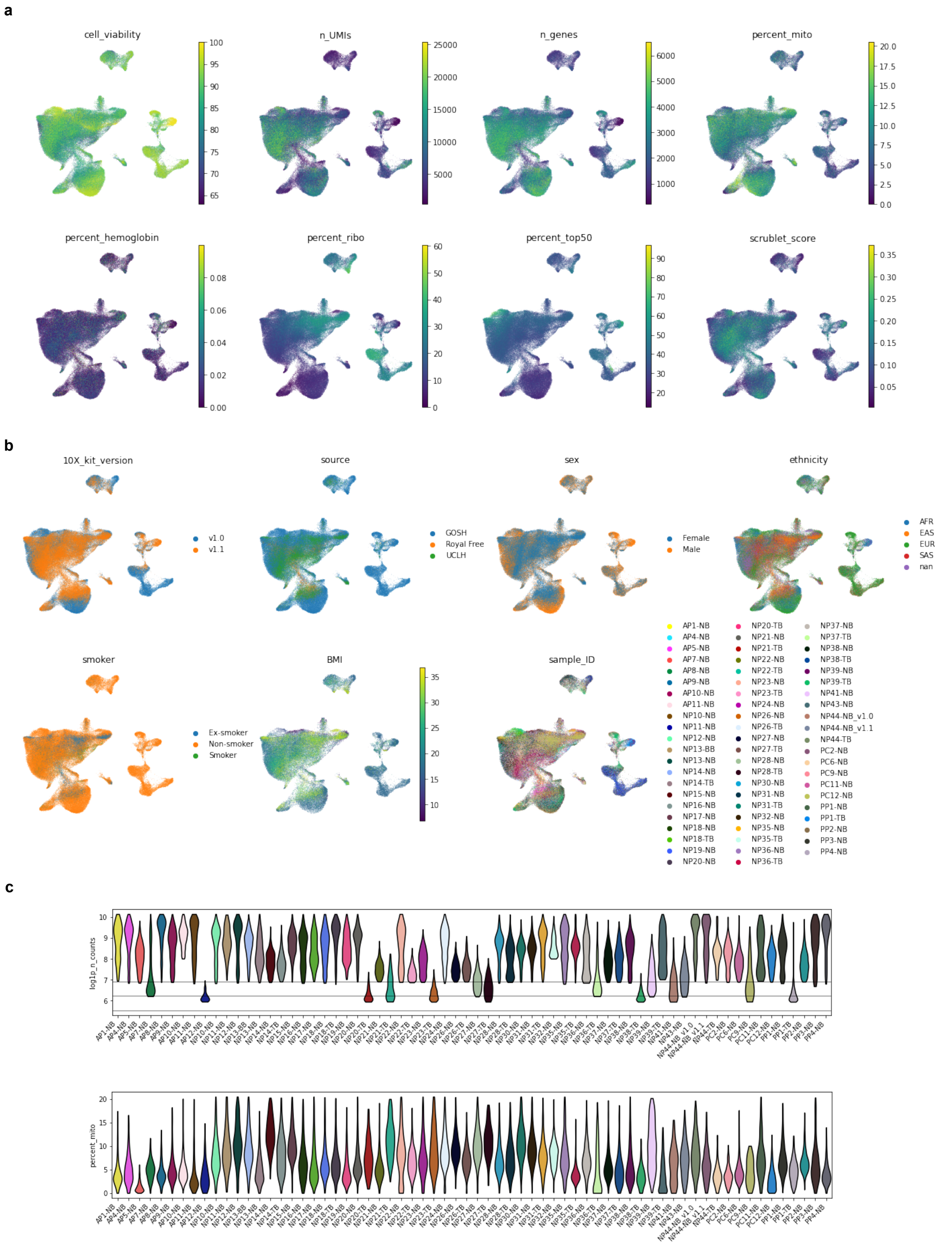

### Extended Data Figure 3

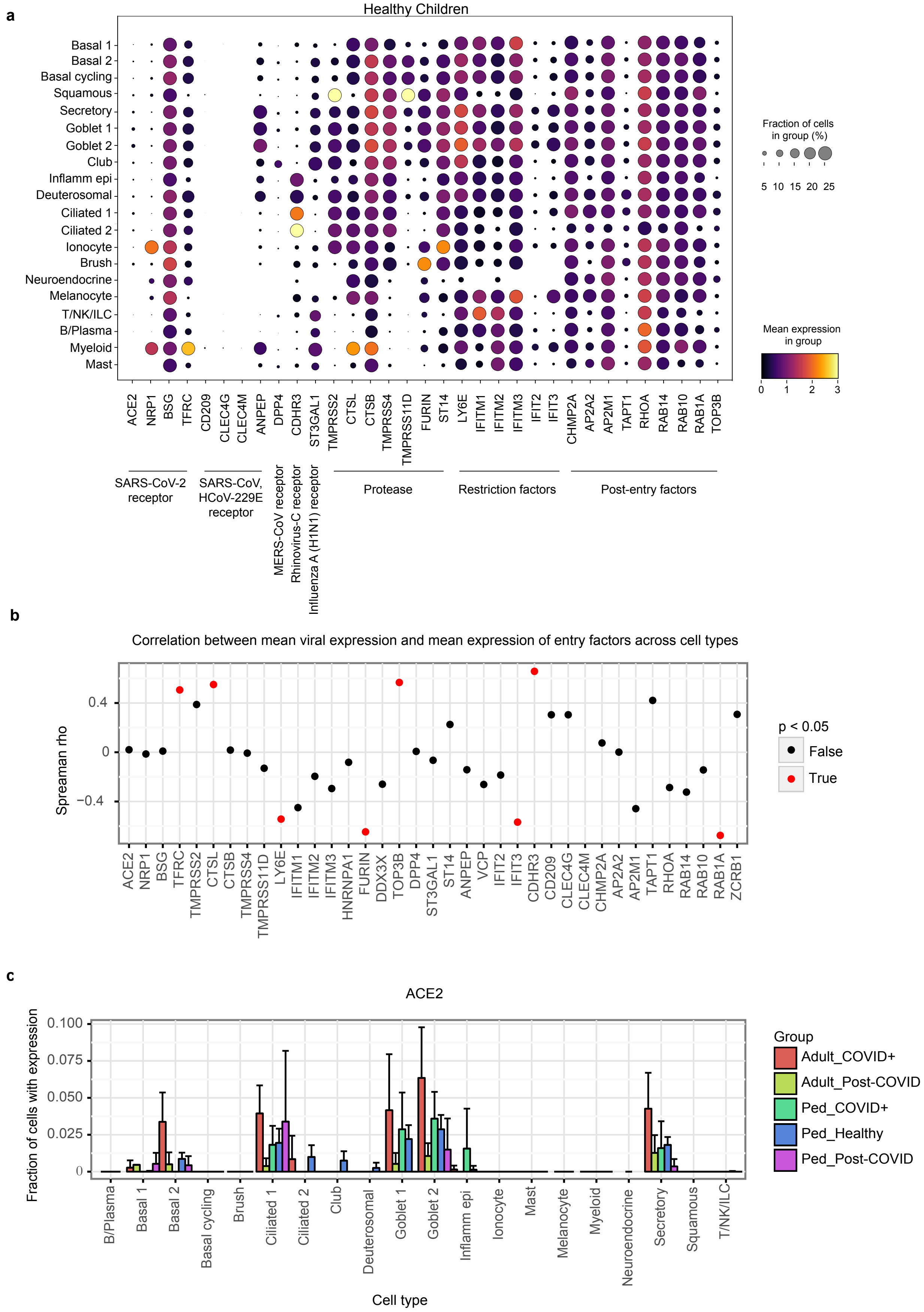

#### Extended Data Figure 4

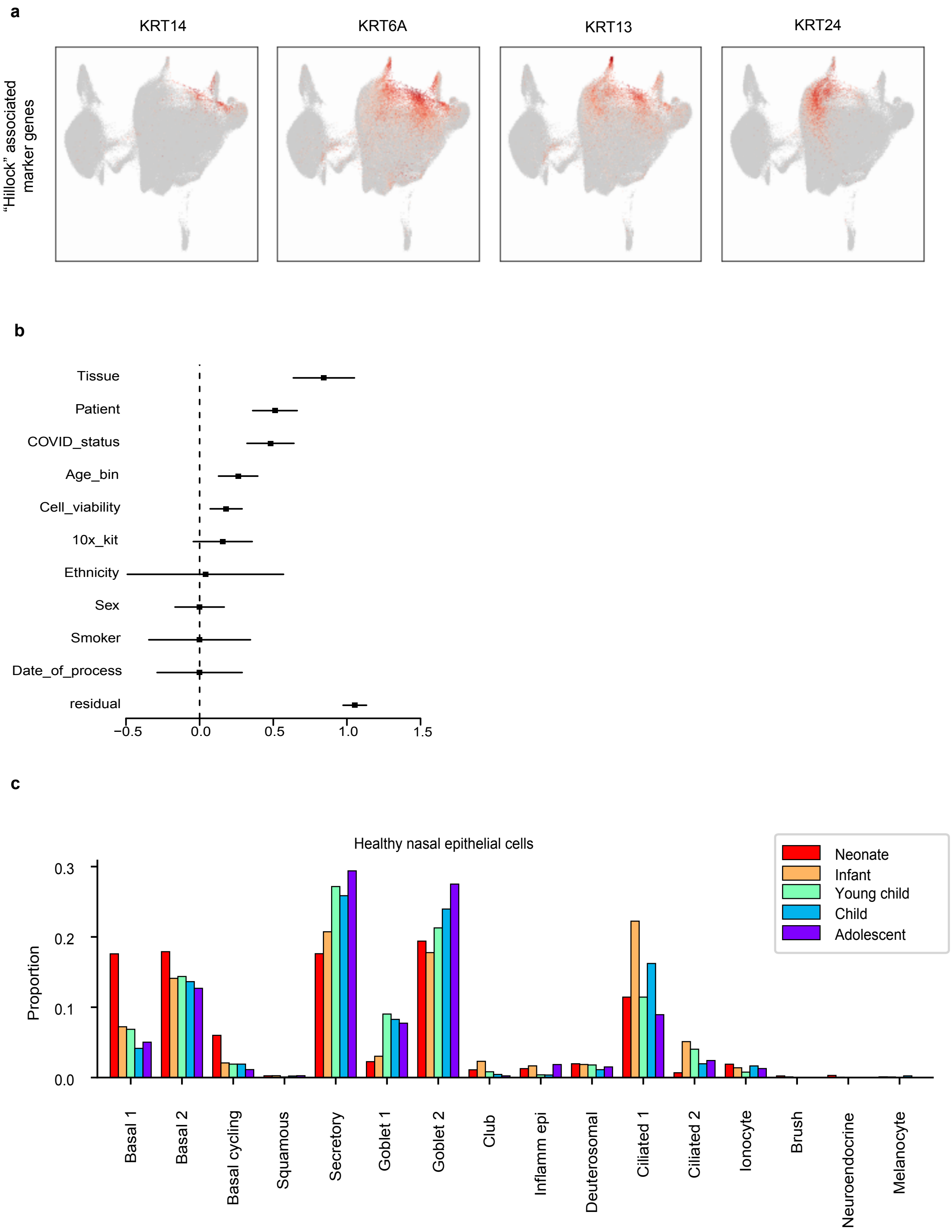

#### Extended Data Figure 5

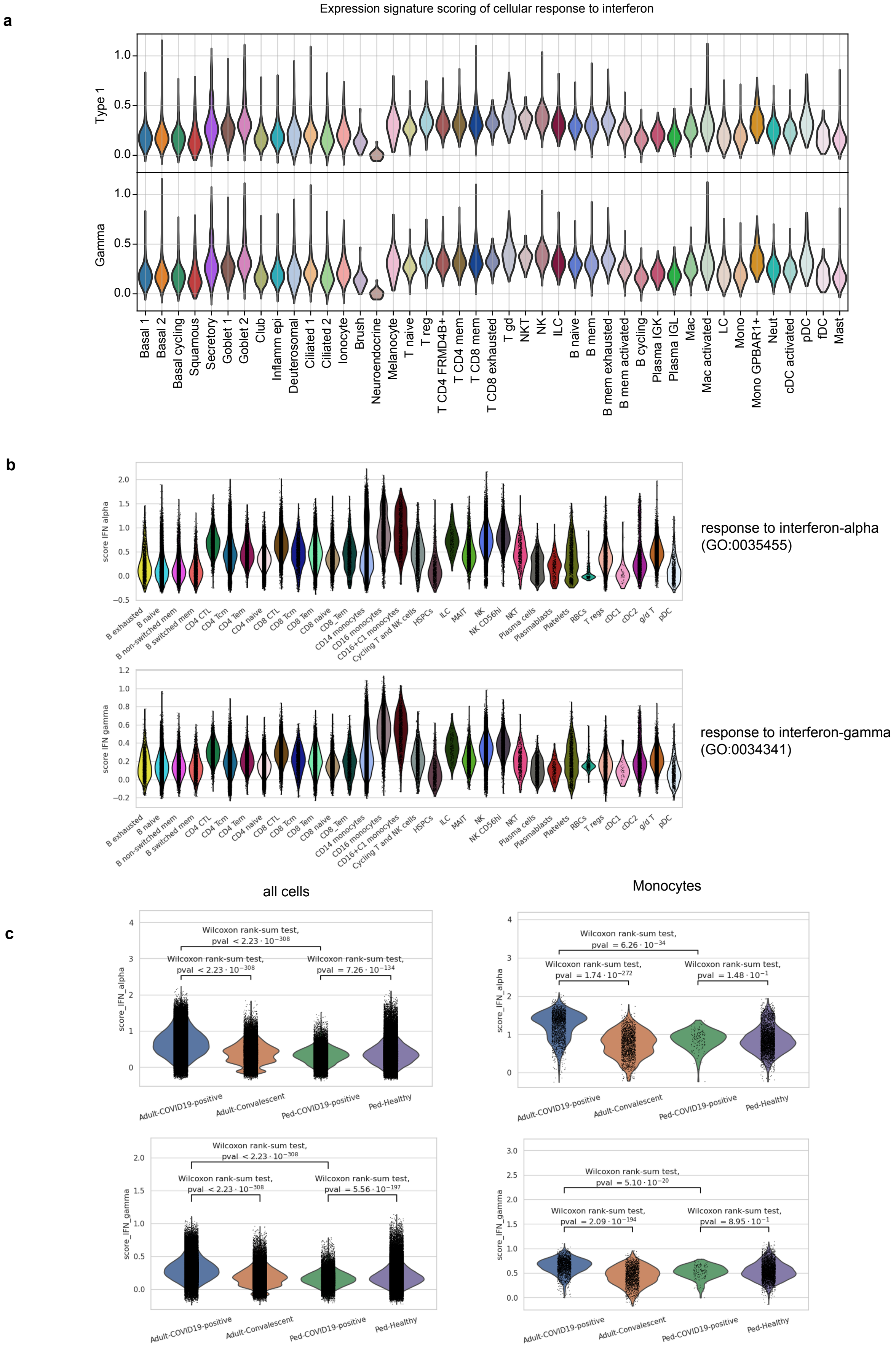

#### Extended Data Figure 6

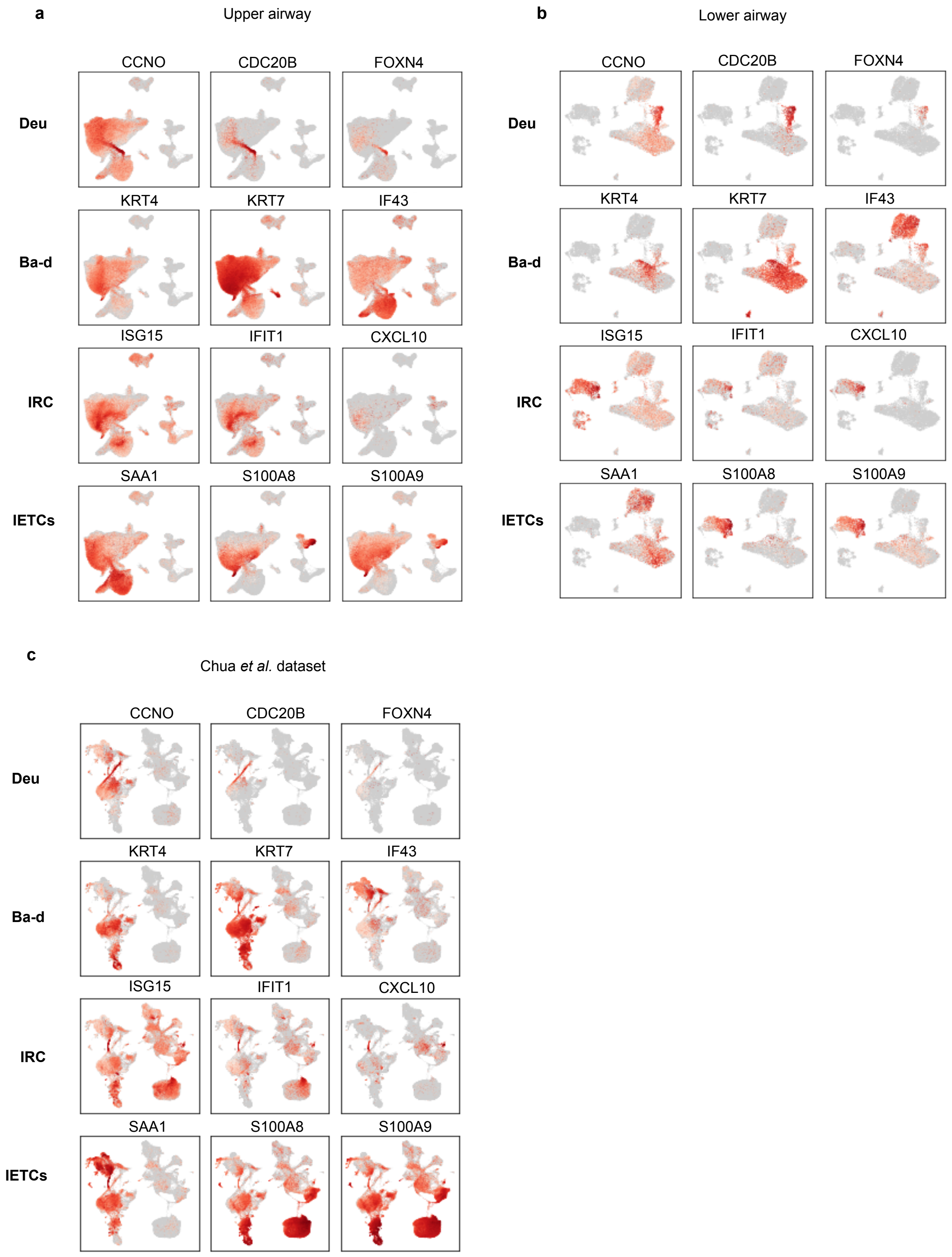

#### Extended Data Figure 7

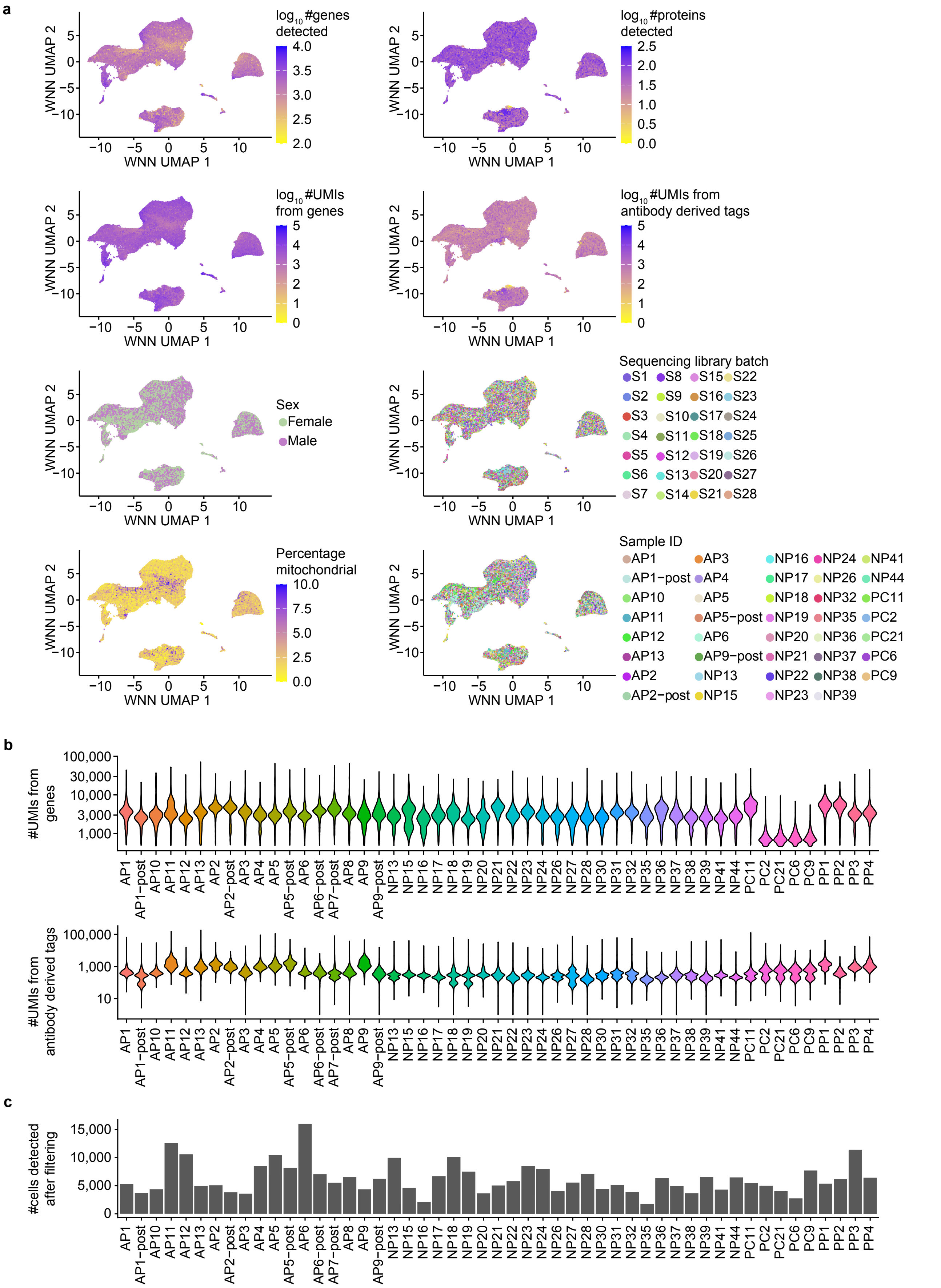

### Extended Data Figure 8

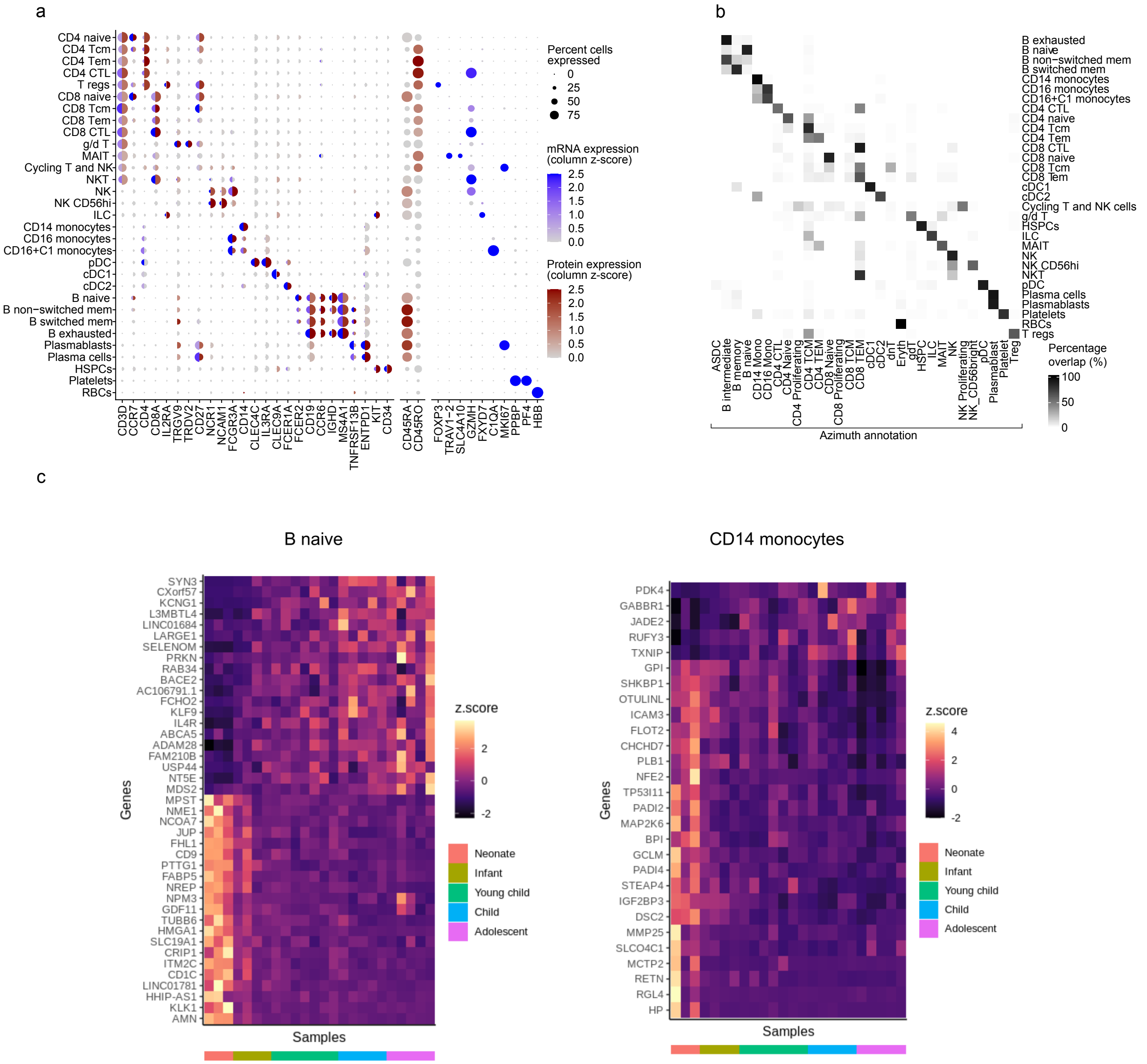

### Extended Data Figure 9

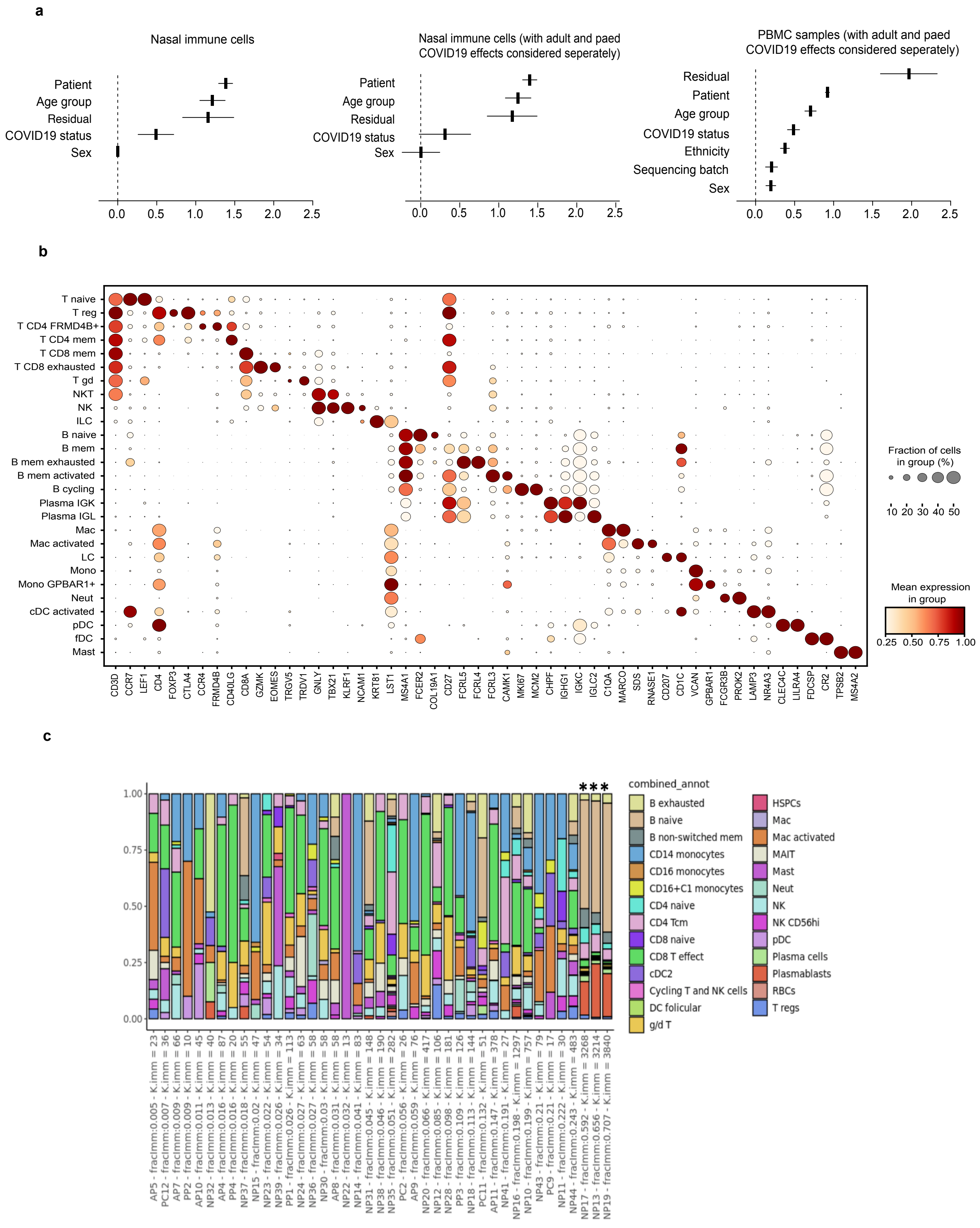

#### Extended Data Figure 10

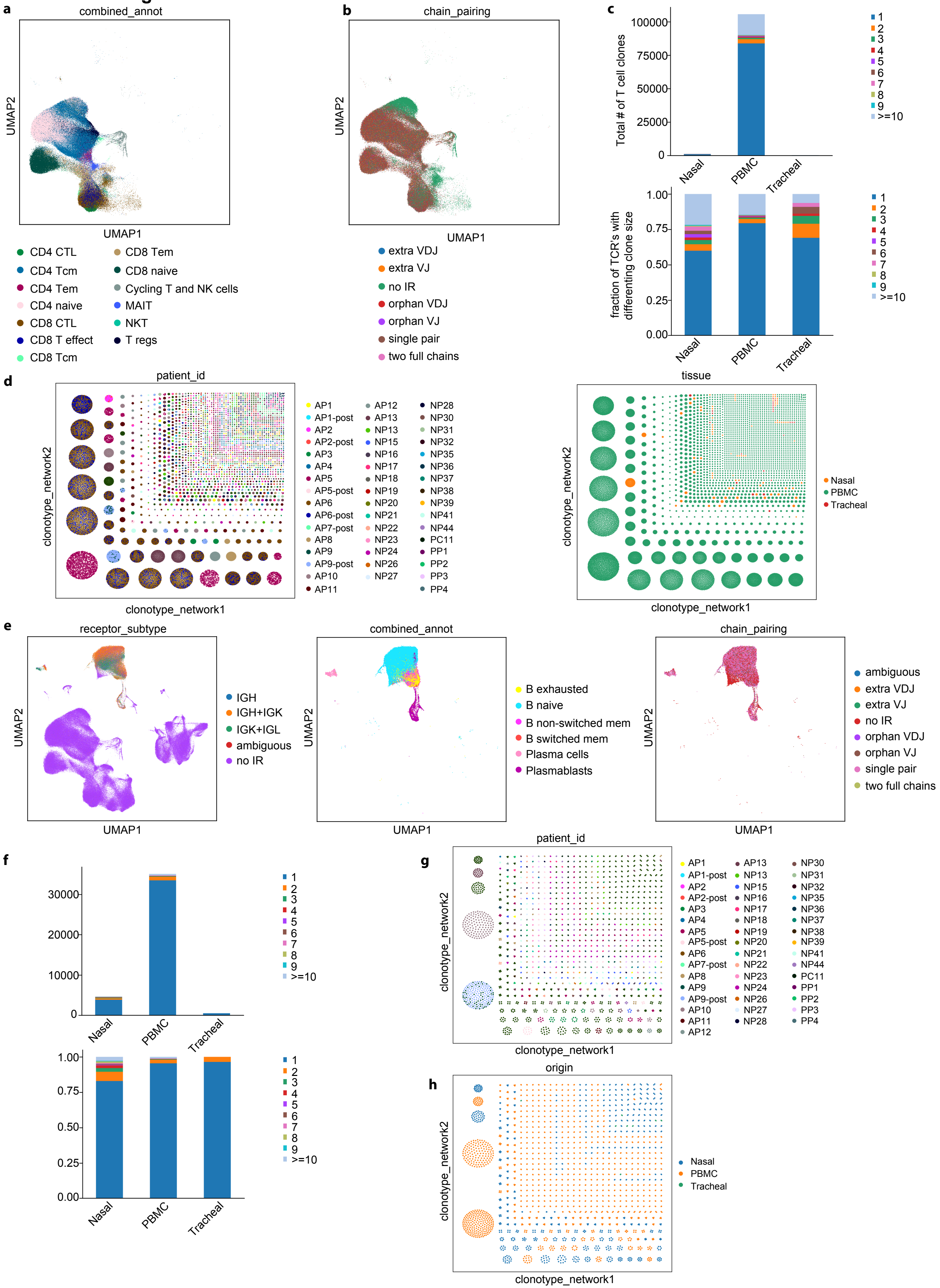

#### Extended Data Table 1

| | SARS-CoV-2 negative children (n = 30) | SARS-CoV-2 positive children (n = 4) | SARS-CoV-2 positive adults (n = 18) | Post-COVID19 (n = 6) |
|---|---|---|---|---|
| **Median age** | 2.87 (3days - 16 yrs) | 5.83 (21 days - 13 yrs) | 66 (25 - 92 yrs) | 35 (4 months - 47 yrs) |
| **Sex** | | | | |
| Male (%) | 13 (43.3) | 3 (75) | 10 (55.6) | 3 (50) |
| Female (%) | 17 (56.7) | 1 (25) | 8 (44.4) | 3 (50) |
| **Ethnicity** | | | | |
| White (%) | 20 (66.7) | 3 (75) | 7 (38.8) | 2 (33.3) |
| Black (%) | 4 (13.3) | 1 (25) | 1 (5.6) | 0 (0) |
| Hispanic (%) | 0 (0) | 0 | 3 (16.7) | 0 (0) |
| South Asian (%) | 1 (3.3) | 0 | 5 (27.7) | 1 (16.7) |
| Other (%) | 2 (6.7) | 0 | 1 (5.6) | 0 (0) |
| Unspecified (%) | 3 (10) | 0 | 1 (5.6) | 3 (50) |
| **Peripheral blood test at sampling** | | | | |
| Lymphocyte count (cells/µl) | 2,026 ± 684.6 | 2,370 ± 1,830 | 1,185 ± 504.1 | 2,160 ± 662.6 |
| **Viral infection symptoms** | | | | |
| Fever, Fatigue (%) | 1 (3.3) | 2 (50) | 13 (72.2) | 3 (50) |
| Digestive symptom (%) | 2 (6.7) | 3 (75) | 13 (72.2) | 2 (33.3) |
| Upper respiratory tract symptom (%) | 3 (10) | 2 (50) | 9 (50) | 3 (50) |
| Respiratory failure (%) | 6 (20) | 4 (100) | 12 (66.7) | 6 (100) |
| **Respiratory Support** | | | | |
| None (%) | - | 0 (0) | 3 (16.7) | 0 (0) |
| Low flow oxygen (%) | - | 0 (0) | 4 (22.2) | 0 (0) |
| HFNC / NIPPV (%) | - | 1 (25) | 4 (22.2) | 3 (50) |
| IMV (%) | - | 3 (75) | 7 (38.9) | 3 (50) |
| **COVID-19 severity** | | | | |
| Asymptomatic (%) | NA | 0 (0) | 0 (0) | 0 (0) |
| Mild (%) | NA | 0 (0) | 5 (27.8) | 0 (0) |
| Moderate (%) | NA | 0 (0) | 3 (16.7) | 0 (0) |
| Severe (%) | NA | 4 (100) | 10 (55.5) | 6 (100) |

Abbreviations: HFNC = high flow nasal canula, NIPPV = non-invasive positive pressure ventilation, IMV = invasive mechanical ventilation, NA = not assesed

### Extended Data Table 2

| DNA_ID | Clone | Description | Barcode |
|---|---|---|---|
| C0005 | 2D10 | 0005 anti-human CD80 | ACGAATCAATCTGTG |
| C0006 | IT2.2 | 0006 anti-human CD86 | GTCTTTGTCAGTGCA |
| C0007 | 29E.2A3 | 0007 anti-human CD274 (B7-H1, PD-L1) | GTTGTCCGACAATAC |
| C0008 | 24F.10C12 | 0008 anti-human CD273 (B7-DC, PD-L2) | TCAACGCTTGGCTAG |
| C0009 | 2D3 | 0009 anti-human CD275 (B7-H2, ICOSL) | GTGCATTCAACAGTA |
| C0014 | M1/70 | 0014 anti-mouse/human CD11b | TGAAGGCTCATTTGT |
| C0021 | 11C3.1 | 0021 anti-human CD252 (OX40L) | TTTAGTGATCCGACT |
| C0022 | 5F4 | 0022 anti-human CD137L (4-1BB Ligand) | ATTCGCCTTACGCAA |
| C0023 | SKII.4 | 0023 anti-human CD155 (PVR) | ATCACATCGTTGCCA |
| C0024 | TX31 | 0024 anti-human CD112 (Nectin-2) | AACCTTCCGTCTAAG |
| C0026 | CC2C6 | 0026 anti-human CD47 | GCATTCTGTCACCTA |
| C0027 | 113-16 | 0027 anti-human CD70 | CGCGAACATAAGAAG |
| C0028 | BY88 | 0028 anti-human CD30 | TCAGGGTGTGCTGTA |
| C0031 | 5C3 | 0031 anti-human CD40 | CTCAGATGGAGTATG |
| C0032 | 24-31 | 0032 anti-human CD154 | GCTAGATAGATGCAA |
| C0033 | HI186 | 0033 anti-human CD52 | CTTTGTACGAGCAAA |
| C0034 | UCHT1 | 0034 anti-human CD3 | CTCATTGTAACTCCT |
| C0046 | SK1 | 0046 anti-human CD8 | GCGCAACTTGATGAT |
| C0047 | 5.1H11 | 0047 anti-human CD56 (NCAM) | TCCTTTCCTGATAGG |
| C0050 | HIB19 | 0050 anti-human CD19 | CTGGGCAATTACTCG |
| C0052 | P67.6 | 0052 anti-human CD33 | TAACTCAGGGCCTAT |
| C0053 | S-HCL-3 | 0053 anti-human CD11c | TACGCCTATAACTTG |
| C0054 | 581 | 0054 anti-human CD34 | GCAGAAATCTCCCTT |
| C0056 | 19F2 | 0056 anti-human CD269 (BCMA) | CAGATGATCCACCAT |
| C0058 | W6/32 | 0058 anti-human HLA-A,B,C | TATGCGAGGCTTATC |
| C0060 | 5E10 | 0060 anti-human CD90 (Thy1) | GCATTGTACGATTCA |
| C0061 | 104D2 | 0061 anti-human CD117 (c-kit) | AGACTAATAGCTGAC |
| C0062 | HI10a | 0062 anti-human CD10 | CAGCCATTCATTAGG |
| C0063 | HI100 | 0063 anti-human CD45RA | TCAATCCTTCCGCTT |
| C0064 | 6H6 | 0064 anti-human CD123 | CTTCACTCTGTCAGG |
| C0066 | CD7-6B7 | 0066 anti-human CD7 | TGGATTCCCGGACTT |
| C0070 | GoH3 | 0070 anti-human/mouse CD49f | TTCCGAGGATGATCT |
| C0071 | L291H4 | 0071 anti-human CD194 (CCR4) | AGCTTACCTGCACGA |
| C0072 | RPA-T4 | 0072 anti-human CD4 | TGTTCCCGCTCAACT |
| C0073 | IM7 | 0073 anti-mouse/human CD44 | TGGCTTCAGGTCCTA |
| C0081 | M5E2 | 0081 anti-human CD14 | TCTCAGACCTCCGTA |
| C0083 | 3G8 | 0083 anti-human CD16 | AAGTTCACTCTTTGC |
| C0085 | BC96 | 0085 anti-human CD25 | TTTGTCCTGTACGCC |
| C0087 | UCHL1 | 0087 anti-human CD45RO | CTCCGAATCATGTTG |
| C0088 | EH12.2H7 | 0088 anti-human CD279 (PD-1) | ACAGCGCCGTATTTA |
| C0089 | A15153G | 0089 anti-human TIGIT (VSTM3) | TTGCTTACCGCCAGA |
| C0090 | MOPC-21 | 0090 Mouse IgG1, κ isotype Ctrl | GCCGGACGACATTAA |
| C0091 | MOPC-173 | 0091 Mouse IgG2a, κ isotype Ctrl | CTCCTACCTAAACTG |
| C0092 | MPC-11 | 0092 Mouse IgG2b, κ isotype Ctrl | ATATGTATCACGCGA |
| C0095 | RTK4530 | 0095 Rat IgG2b, κ Isotype Ctrl | GATTCTTGACGACCT |
| C0100 | 2H7 | 0100 anti-human CD20 | TTCTGGGTCCCTAGA |
| C0101 | 9E2 | 0101 anti-human CD335 (NKp46) | ACAATTTGAACAGCG |
| C0102 | BM16 | 0102 anti-human CD294 (CRTH2) | TGTTTACGAGAGCCC |
| C0123 | 9C4 | 0123 anti-human CD326 (Ep-CAM) | TTCCGAGCAAGTATC |
| C0124 | WM59 | 0124 anti-human CD31 | ACCTTTATGCCACGG |
| C0127 | NC-08 | 0127 anti-Human Podoplanin | GGTTACTCGTTGTGT |
| C0134 | P1H12 | 0134 anti-human CD146 | CCTTGGATAACATCA |
| C0135 | 67A4 | 0135 anti-human CD324 (E-Cadherin) | ATCCTTCTCCCTTTC |
| C0136 | MHM-88 | 0136 anti-human IgM | TAGCGAGCCCGTATA |
| C0138 | UCHT2 | 0138 anti-human CD5 | CATTAACGGGATGCC |
| C0139 | B1 | 0139 anti-human TCR γ/δ | CTTCCGATTCATTCA |
| C0140 | G025H7 | 0140 anti-human CD183 (CXCR3) | GCGATGGTAGATTAT |
| C0141 | J418F1 | 0141 anti-human CD195 (CCR5) | CCAAAGTAAGAGCCA |
| C0142 | FUN-2 | 0142 anti-human CD32 | GCTTCCGAATTACCG |
| C0143 | G034E3 | 0143 anti-human CD196 (CCR6) | GATCCCTTTGTCACT |
| C0144 | J252D4 | 0144 anti-human CD185 (CXCR5) | AATTCAACCGTCGCC |
| C0145 | Ber-ACT8 | 0145 anti-human CD103 (Integrin αE) | GACCTCATTGTGAAT |
| C0146 | FN50 | 0146 anti-human CD69 | GTCTCTTGGCTTAAA |
| C0147 | DREG-56 | 0147 anti-human CD62L | GTCCCTGCAACTTGA |
| C0148 | G043H7 | 0148 anti-human CD197 (CCR7) | AGTTCAGTCAACCGA |
| C0149 | HP-3G10 | 0149 anti-human CD161 | GTACGCAGTCCTTCT |
| C0151 | BNI3 | 0151 anti-human CD152 (CTLA-4) | ATGGTTCACGTAATC |
| C0152 | 11C3C65 | 0152 anti-human CD223 (LAG-3) | CATTTGTCTGCCGGT |
| C0153 | SA231A2 | 0153 anti-human KLRG1 (MAFA) | CTTATTTCCTGCCCT |
| C0154 | O323 | 0154 anti-human CD27 | GCACTCCTGCATGTA |
| C0155 | H4A3 | 0155 anti-human CD107a (LAMP-1) | CAGCCCACTGCAATA |
| C0156 | DX2 | 0156 anti-human CD95 (Fas) | CCAGCTCATTAGAGC |
| C0159 | L243 | 0159 anti-human HLA-DR | AATAGCGAGCAAGTA |
| C0160 | L161 | 0160 anti-human CD1c | GAGCTACTTCACTCG |
| C0162 | 10.1 | 0162 anti-human CD64 | AAGTATGCCCTACGA |
| C0163 | M80 | 0163 anti-human CD141 (Thrombomodulin) | GGATAACCGCGCTTT |
| C0164 | 51.1 | 0164 anti-human CD1d | TCGAGTCGCTTATCA |
| C0165 | 1D11 | 0165 anti-human CD314 (NKG2D) | CGTGTTTGTTCCTCA |
| C0166 | 6/40c | 0166 anti-human CD66b | AGCTGTAAGTTTCGG |
| C0167 | E11 | 0167 anti-human CD35 | ACTTCCGTCGATCTT |
| C0168 | QA17A04 | 0168 anti-human CD57 Recombinant | AACTCCCTATGGAGG |
| C0169 | F38-2E2 | 0169 anti-human CD366 (Tim-3) | TGTCCTACCCAACTT |
| C0170 | MIH26 | 0170 anti-human CD272 (BTLA) | GTTATTGGACTAAGG |
| C0171 | C398.4A | 0171 anti-human/mouse/rat CD278 (ICOS) | CGCGCACCCATTAAA |
| C0174 | TS2/9 | 0174 anti-human CD58 (LFA-3) | GTTCCTATGGACGAC |
| C0175 | NK92.39 | 0175 anti-human CD96 (TACTILE) | TGGCCTATAAATGGT |
| C0176 | A1 | 0176 anti-human CD39 | TTACCTGGTATCCGT |
| C0177 | NOK-1 | 0177 anti-human CD178 (Fas-L) | CCGGTCCTCTGTATT |
| C0179 | K0124E1 | 0179 anti-human CX3CR1 | AGTATCGTCTCTGGG |
| C0180 | ML5 | 0180 anti-human CD24 | AGATTCCTTCGTGTT |
| C0181 | Bu32 | 0181 anti-human CD21 | AACCTAGTAGTTCGG |
| C0185 | TS2/4 | 0185 anti-human CD11a | TATATCCTTGTGAGC |
| C0186 | HP6123 | 0186 anti-human IgA | AAGATGTCCGAGCAA |
| C0187 | CB3-1 | 0187 anti-human CD79b (Igβ) | ATTCTTCAACCGAAG |
| C0188 | ASL-32 | 0188 anti-human CD66a/c/e | GGGACAGTTCGTTTC |
| C0189 | C1.7 | 0189 anti-human CD244 (2B4) | TCGCTTGGATGGTAG |

| DNA_ID | Clone | Description | Barcode |
|---|---|---|---|
| C0196 | HIR2 | 0196 anti-human CD235ab | GCTCCTTTACACGTA |
| C0205 | 15-2 | 0205 anti-human CD206 (MMR) | TCAGAACGTCTAACT |
| C0206 | 7-239 | 0206 anti-human CD169 (Sialoadhesin, Siglec-1) | TACTCAGCGTGTTTG |
| C0207 | 8F9 | 0207 anti-human CD370 (CLEC9A/DNGR1) | CTGCATTTCAGTAAG |
| C0208 | S15046E | 0208 anti-human XCR1 | AAGACGCATGTCAAC |
| C0214 | FIB504 | 0214 anti-human/mouse integrin β7 | TCCTTGGATGTACCG |
| C0215 | 11C1 | 0215 anti-human CD268 (BAFF-R) | CGAAGTCGATCCGTA |
| C0217 | HA58 | 0217 anti-human CD54 | CTGATAGACTTGAGT |
| C0218 | AK4 | 0218 anti-human CD62P (P-Selectin) | CCTTCCGTATCCCTT |
| C0224 | IP26 | 0224 anti-human TCR α/β | CGTAACGTAGAGCGA |
| C0245 | STA | 0245 anti-human CD106 | TCACAGTTCCTTGGA |
| C0246 | TU27 | 0246 anti-human CD122 (IL-2Rβ) | TCATTTCCTCCGATT |
| C0247 | 1A1 | 0247 anti-human CD267 (TACI) | AGTGATGGAGCGAAC |
| C0352 | AER-37 (CRA-1) | 0352 anti-human Fc εRIα | CTCGTTTCCGTATCG |
| C0353 | HIP8 | 0353 anti-human CD41 | ACGTTGTGGCCTTGT |
| C0355 | 4B4-1 | 0355 anti-human CD137 (4-1BB) | CAGTAAGTTCGGGAC |
| C0356 | MIH24 | 0356 anti-human CD254 (TRANCE, RANKL) | TCCGTGTTAGTTTGT |
| C0358 | GHI/61 | 0358 anti-human CD163 | GCTTCTCCTTCCTTA |
| C0359 | HB15e | 0359 anti-human CD83 | CCACTCATTTCCGGT |
| C0360 | 108-17 | 0360 anti-human CD357 (GITR) | ACCTTTCGACACTCG |
| C0362 | 7D4-6 | 0362 anti-human CD309 (VEGFR2) | TTCACGCAGTAAGAT |
| C0363 | G077F6 | 0363 anti-human CD124 (IL-4Rα) | CCGTCCTGATAGATG |
| C0366 | 12G5 | 0366 anti-human CD184 (CXCR4) | TCAGGTCCTTTCAAC |
| C0367 | TS1/8 | 0367 anti-human CD2 | TACGATTTGTCAGGG |
| C0368 | 11A8 | 0368 anti-human CD226 (DNAM-1) | TCTCAGTGTTTGTGG |
| C0369 | TS2/16 | 0369 anti-human CD29 | GTATTCCCTCAGTCA |
| C0370 | 201A | 0370 anti-human CD303 (BDCA-2) | GAGATGTCCGAATTT |
| C0371 | P1E6-C5 | 0371 anti-human CD49b | GCTTTCTTCAGTATG |
| C0373 | 5A6 | 0373 anti-human CD81 (TAPA-1) | GTATCCTTCCTTGGC |
| C0374 | MEM-108 | 0374 anti-human CD98 | GCACCAACAGCCATT |
| C0375 | M1310G05 | 0375 anti-human IgG Fc | CTGGAGCGATTAGAA |
| C0384 | IA6-2 | 0384 anti-human IgD | CAGTCTCCGTAGAGT |
| C0385 | TS1/18 | 0385 anti-human CD18 | TATTGGGACACTTCT |
| C0386 | CD28.2 | 0386 anti-human CD28 | TGAGAACGACCCTAA |
| C0387 | 1D3 | 0387 anti-human TSLPR (TSLP-R) | CAGTCCTCTCTGTCA |
| C0389 | HIT2 | 0389 anti-human CD38 | TGTACCCGCTTGTGA |
| C0390 | A019D5 | 0390 anti-human CD127 (IL-7Rα) | GTGTGTTGTCCTATG |
| C0391 | HI30 | 0391 anti-human CD45 | TGCAATTACCCGGAT |
| C0392 | W6D3 | 0392 anti-human CD15 (SSEA-1) | TCACCAGTACCTAGT |
| C0393 | S-HCL-1 | 0393 anti-human CD22 | GGGTTGTTGTCTTTG |
| C0394 | CY1G4 | 0394 anti-human CD71 | CCGTGTTCCTCATTA |
| C0395 | MIH43 | 0395 anti-human B7-H4 | TGTATGTCTGCCTTG |
| C0396 | BA5b | 0396 anti-human CD26 | GGTGGCTAGATAATG |
| C0397 | 5E8 | 0397 anti-human CD193 (CCR3) | ACCAATCCTTTCGTC |
| C0399 | 7C9C20 | 0399 anti-human CD204 | TAGCGAGCCAGATGT |
| C0400 | BV9 | 0400 anti-human CD144 (VE-Cadherin) | TCCACTCATTCTGTA |
| C0402 | HI149 | 0402 anti-human CD1a | GATCGTGTTGTGTTA |
| C0406 | 12C2 | 0406 anti-human CD304 (Neuropilin-1) | GGACTAAGTTTCGTT |
| C0407 | 5-271 | 0407 anti-human CD36 | TTCTTTGCCTTGCCA |
| C0420 | HP-MA4 | 0420 anti-human CD158 (KIR2DL1/S1/S3/S5) | TATCAACCAACGCTT |
| C0437 | 4C7 | 0437 anti-mouse/human CD207 | CGATTTGTATTCCCT |
| C0576 | 9F10 | 0576 anti-human CD49d | CCATTCAACTTCCGG |
| C0577 | AD2 | 0577 anti-human CD73 (Ecto-5'-nucleotidase) | CAGTTCCTCAGTTCG |
| C0581 | 3C10 | 0581 anti-human TCR Vα7.2 | TACGAGCAGTATTCA |
| C0582 | B6 | 0582 anti-human TCR Vδ2 | TCAGTCAGATGGTAT |
| C0583 | B3 | 0583 anti-human TCR Vγ9 | AAGTGATGGTATCTG |
| C0584 | 6B11 | 0584 anti-human TCR Vα24-Jα18 (iNKT cell) | AACTTCTGTGGTAGC |
| C0590 | NKTA255 | 0590 anti-human CD305 (LAIR1) | ATTTCCATTCCCTGT |
| C0591 | 15C4 | 0591 anti-human LOX-1 | ACCCTTTACCGAATA |
| C0592 | DX27 | 0592 anti-human CD158b (KIR2DL2/L3, NKAT2) | GACCCGTAGTTTGAT |
| C0594 | S16016B | 0594 anti-human CD133 | GTAAGACGCCTATGC |
| C0597 | 9E9A8 | 0597 anti-human CD209 (DC-SIGN) | TCACTGGACACTTAA |
| C0599 | DX9 | 0599 anti-human CD158e1 (KIR3DL1, NKB1) | GGACGCTTTCCTTGA |
| C0600 | UP-R1 | 0600 anti-human CD158f (KIR2DL5) | AAAGTGATGCCACTG |
| C0801 | P30-15 | 0801 anti-human CD337 (NKp30) | AAAGTCACTCTGCCG |
| C0802 | P44-8 | 0802 anti-human CD336 (NKp44) | GGGCAATTAGCGAGT |
| C0828 | 413D12 | 0828 anti-human CD307d (FcRL4) | CGATTTGATCTGCCT |
| C0829 | 509f6 | 0829 anti-human CD307e (FcRL5) | TCACGCAGTCCTCAA |
| C0830 | 162.1 | 0830 anti-human CD319 (CRACC) | AGTATGCCATGTCTT |
| C0831 | DL-101 | 0831 anti-human CD138 (Syndecan-1) | GTATAGACCAAAGCC |
| C0845 | 3B2/TA8 | 0845 anti-human CD99 | ACCCGTCCCTAAGAA |
| C0853 | 50C1 | 0853 anti-human CLEC12A | CATTAGAGTCTGCCA |
| C0856 | M7004D06 | 0856 anti-Tau Phospho (Thr181) | CTCGTTTGTAGCAAT |
| C0863 | 1D6 | 0863 anti-human CD257 (BAFF, BLYS) | CAGAGCACCCATTAA |
| C0867 | DX22 | 0867 anti-human CD94 | CTTTCCGGTCCTACA |
| C0870 | A12 (7D4) | 0870 anti-human CD150 (SLAM) | GTCATTGTATGTCTG |
| C0894 | MHK-49 | 0894 anti-human Ig light chain κ | AGCTCAGCCAGTATG |
| C0895 | M3/38 | 0895 anti-mouse/human Mac-2 (Galectin-3) | GATGCAATTAGCCGG |
| C0896 | GHI/75 | 0896 anti-human CD85j (ILT2) | CCTTGTGAGGCTATG |
| C0897 | EBVCS-5 | 0897 anti-human CD23 | TCTGTATAACCGTCT |
| C0898 | MHL-38 | 0898 anti-human Ig light chain λ | CAGCCAGTAAGTCAC |
| C0899 | BB7.2 | 0899 anti-human HLA-A2 | GAACATTTCCGACAA |
| C0901 | 7B11 | 0901 anti-human GARP (LRRC32) | AGGTATGGTAGAGTA |
| C0902 | 6-434 | 0902 anti-human CD328 (Siglec-7) | CTTAGCATTTCACTG |
| C0908 | H131 | 0908 anti-human TCR Vβ13.1 | TTATGGACGTATGGT |
| C0920 | ASL-24 | 0920 anti-human CD82 | TCCCACTTCCGCTTT |
| C0944 | BB27 | 0944 anti-human CD101 (BB27) | CTACTTCCCTGTCAA |
| C0985 | 4B2.9 | 0985 anti-human CD360 (IL-21R) | GAGGATGATGCCATG |
| C1046 | S5/1 | 1046 anti-human CD88 (C5aR) | GCCGCATGAGAAACA |
| C1047 | 3D11/HLA-F | 1047 anti-human HLA-F | GCAACTCTCCTACCT |
| C1048 | 8F10B51 | 1048 anti-human NLRP2 | ACGCTTGTGTTAGTT |
| C1051 | mAb 84 | 1051 anti-human Podocalyxin | GAGCCGGTATAATGC |
| C1052 | KF29 | 1052 anti-human CD224 | CTGATGAGATGTCAG |
| C1055 | 12.1 | 1055 anti-c-Met | GCTGCTGCGATTTGA |
| C1056 | T5-39 | 1056 anti-human CD258 (LIGHT) | ACTTCCCTGTAGAAA |
| C1057 | JD3 | 1057 anti-human DR3 (TRAMP) | GAGTTCCCTCAGTTC |

### Extended Data Table 3

| Virus_id | Virus name | link |
|---|---|---|
| NC_007605.1 | EBV1 (Human gammaherpesvirus 4) | https://www.ncbi.nlm.nih.gov/nuccore/82503188 |
| NC_009334.1 | EBV2 (Human herpesvirus 4) | https://www.ncbi.nlm.nih.gov/nuccore/139424470 |
| AF156963 | ERVWE1 (syncytin) | https://www.ncbi.nlm.nih.gov/nuccore/AF156963 |
| AY101582 | ERVWE1 (syncytin) | https://www.ncbi.nlm.nih.gov/nuccore/AY101582 |
| AY101583 | ERVWE1 (syncytin) | https://www.ncbi.nlm.nih.gov/nuccore/AY101583 |
| AY101584 | ERVWE1 (syncytin) | https://www.ncbi.nlm.nih.gov/nuccore/AY101584 |
| AY101585 | ERVWE1 (syncytin) | https://www.ncbi.nlm.nih.gov/nuccore/AY101585 |
| AF072498 | HERV-W | https://www.ncbi.nlm.nih.gov/nuccore/AF072498 |
| AF127228 | HERV-W | https://www.ncbi.nlm.nih.gov/nuccore/AF127228 |
| AF127229 | HERV-W | https://www.ncbi.nlm.nih.gov/nuccore/AF127229 |
| AF331500 | HERV-W | https://www.ncbi.nlm.nih.gov/nuccore/AF331500.1/ |
| NC_001664.4 | HHV-6A (Human Herpes Virus 6) | https://www.ncbi.nlm.nih.gov/nuccore/1344462938 |
| NC_000898.1 | HHV-6B (Human Herpes Virus 6) | https://www.ncbi.nlm.nih.gov/nuccore/9633069 |
| NC_001806.2 | Herpes Simplex Virus 1 (Human alphaherpesvirus 1) | https://www.ncbi.nlm.nih.gov/nuccore/820945227 |
| NC_001798.2 | Herpes Simplex Virus 2 (Human alphaherpesvirus 1) | https://www.ncbi.nlm.nih.gov/nuccore/820945149 |
| NC_001498.1 | Measles morbillivirus | https://www.ncbi.nlm.nih.gov/nuccore/9626945 |
| NC_002200.1 | Mumps rubulavirus | https://www.ncbi.nlm.nih.gov/nuccore/9695415 |
| NC_001545.2 | Rubella | https://www.ncbi.nlm.nih.gov/nuccore/336284682 |
| NC_001348.1 | Varicella Zoster Virus (VZV Human alphaherpesvirus 3) | https://www.ncbi.nlm.nih.gov/nuccore/9625875 |
| NC_006273.2 | Cytomegalovieus (CMV) | https://www.ncbi.nlm.nih.gov/nuccore/155573622 |
| NC_045512.2 | SARS-CoV2 | https://www.ncbi.nlm.nih.gov/nuccore/1798174254 |
